# Impact of Detection-Isolation-Leakage on the 2026 DRC Bundibugyo Ebolavirus Outbreak

**DOI:** 10.64898/2026.08.25.26361360

**Authors:** Tamer Oraby, Dadi Falay, Martial L. Ndeffo-Mbah

## Abstract

The 17th Ebola outbreak in the Democratic Republic of the Congo, announced on 15 May 2026, was attributed to *Bundibugyo ebolavirus* (BDBV). Although case isolation is the main control strategy, its effectiveness is compromised when patients escape isolation facilities before recovery. Between 14 May and 17 June 2026, 175 individuals reportedly left isolation facilities without formal discharge across Ituri Province. We assessed how this “isolation leakage” affects community transmission.

We refined the SEIHFR framework to distinguish undetected community infections, detected but not-yet-isolated cases, isolated individuals, leakage, funeral-associated transmission, and removals. Using Bayesian inference, we fitted the model to daily Ituri surveillance data, escapee counts, and isolation census records. We estimated the leakage rate, reporting and detection probabilities, and the transmission rate, while fixing other parameters based on the BDBV literature.

The model reproduced confirmed cases, deaths, discharges, and escapees. We estimated *R*_0_ = 3.67 (95% HDI: 2.0–5.7), a leakage rate of *ρ ≈* 0.034 day*^−^*^1^ (0.022–0.051), and high contact-tracing-driven detection (*p_d_ ≈* 0.91–0.99). Leakage increased the detection-dependent reproduction number *R*(*p_d_*) from approximately 3.2 to above 5. Eliminating leakage reduced cumulative infections by about one-third, from 1,120 to 764, while the minimum detection level required for control increased from *p_d_ ≥* 0.73 without leakage to *p_d_ ≥* 0.87 at the fitted leakage rate. Shortening time to isolation prevented the most infections (73.4%; 59–84), followed by reducing leakage (29.7%; 14–52) and re-isolating escapees (12.6%; 6–24). Delaying leakage reduction until week 4 reduced its benefit from about 27% to below 2%.

Isolation leakage represents a major transmission pathway that has until now gone largely unmeasured. While rapid initiation of isolation is highly beneficial, it cannot compensate for permeable isolation; therefore, early, community-driven efforts to control leakage, embedded within a multilayered response, are critical.

**Author Summary:** In 2026, an Ebola outbreak caused by the Bundibugyo virus emerged in the northeastern Democratic Republic of the Congo. Since there is no vaccine or approved treatment for this strain, health workers must depend on rapid case detection, prompt isolation, and safe burials to halt transmission. However, surveillance data highlighted a persistent challenge: many patients escaped isolation centers before fully recovering and returned to their communities while still contagious. We refer to this as “isolation leakage.” Although frequently reported during Ebola outbreaks, it has rarely been analyzed using mathematical modeling. We developed a model that follows undetected infections in the community, patients in isolation, and those who escape isolation, and calibrated it to daily surveillance data from Ituri Province, including daily counts of people who escaped Ebola isolation centers. Our analysis showed that leakage significantly amplified the size of the outbreak, and that early action was crucial; intervening to reduce leakage in the first week averted far more infections than waiting until a month into the outbreak. Shortening the time to isolation was the most effective single intervention. These findings indicate that ensuring patients remain in care is vital for controlling the 2026 Ebola outbreak in the fragile, conflict-affected region of Eastern DRC.

## Introduction

Ebola disease is a zoonotic viral illness caused by pathogenic members of the genus *Orthoebola virus*.^15,22^ Human outbreaks occur when the virus is transmitted from animal reservoirs or intermediate hosts to humans, enabling human-to-human transmission that sustains local or regional outbreaks.^15,34^ The primary human disease-causing ebolaviruses include Zaire ebolavirus (EBOV), Sudan ebolavirus (SUDV), and Bundibugyo ebolavirus (BDBV).^15,28^ The other three species, Taï Forest, Reston, and Bombali ebolavirus, have never caused a human outbreak.^15,28,41^ This distinction is crucial for outbreak management because no licensed vaccine covers all ebolavirus species.^19,21^

The 17th Ebola epidemic in the DRC was officially declared on May 15, 2026, after genomic sequencing confirmed Bundibugyo ebolavirus as the cause.^5,38^ The outbreak began in the Mongbwalu health zone of Ituri Province in northeastern DRC and, by 30 May 2026, had expanded to 22 health zones across three provinces (Ituri, Nord-Kivu, and Sud-Kivu).^11^ As of that date, there were 282 confirmed cases and 42 confirmed deaths, yielding a case fatality ratio of 14.9%. Active transmission was ongoing in 14 Ituri health zones. This is only the third documented BDBV outbreak, after the 2007 Uganda outbreak and the 2012 DRC Isiro outbreak, and by far the largest.^32,17,6^

Ebolavirus transmission occurs through direct contact with the bodily fluids of infected individuals and with contaminated items (fomites). These include blood, vomit, diarrhea, sweat, saliva, semen, breast milk, contaminated items, healthcare exposures, and deceased bodies.^39^ The incubation period typically ranges from 2 to 21 days, with an average of 8 to 10 days.^39,8^ People are typically not contagious before symptoms appear. This transmission pattern, along with increased healthcare- and funeral-related exposures, makes the traditional SEIHFR (Susceptible-Exposed-Infectious-Hospitalized-Funeral-Recovered) model—distinguishing community, healthcare, and funeral transmission pathways—the most suitable framework for analyzing Ebola outbreaks.^13,12^

A key but often overlooked aspect of real-world Ebola response, especially in conflict zones and low-trust environments, is *isolation leakage*. This occurs when patients leave healthcare or isolation facilities before fully recovering and return to the community, where they can still transmit the virus. Isolation leakage has been an ongoing challenge for the Ebola response in the DRC. For example, in the Bunia health-zone cluster, escapees (*évadés*) were recorded on every day of observation from 20 May to 1 June 2026, with a total of 60 leakage events across 13 days at 13 different facilities.^31^ The highest number of escapees in a single day was 10, recorded on 28 May and 30 May. This indicates a structural transmission pathway that, as far as we know, has not been explicitly investigated in previous modeling studies, despite being well documented during previous Ebola outbreaks in the DRC.

The current large-scale Bundibugyo virus (BDBV) outbreak in the DRC exhibits several distinguishing features compared with recent outbreaks in the country. Currently, there is no licensed vaccine for BDBV in such an emergency. The rVSV-ZEBOV vaccine (Ervebo) provides protection against EBOV but does not confer immunity to BDBV.^42^ Moreover, the monoclonal antibody therapies approved for EBOV, such as mAb114 and REGN-EB3, are not efficacious against BDBV.^43,44^ The observed case fatality rate of approximately 14.9% in the 2026 outbreak is consistent with the 25-36% range reported during the 2007 Uganda BDBV outbreak and is lower than the typical 40-90% observed in EBOV outbreaks.^32,17,14^

A systematic review^24^ found that approximately 92% of Ebola virus disease (EVD) models in the literature focus on the Zaire species. The new model introduces a leakage compartment and transmission dynamics.^24^ Additionally, the Ituri province in DRC, the epicenter of the ongoing BDBV outbreak, is marked by a persistent humanitarian crisis due to the ongoing armed conflict, resulting in over one million internally displaced persons and a severely strained health system. These factors increase healthcare worker exposure risk due to shortages of personal protective equipment and create substantial operational barriers to effective contact tracing.^10,11^

This paper examines how much additional community transmission occurs when patients leave Ebola treatment or isolation centers before completing their treatment, especially when some infections go undetected. The main innovation is the addition of a leakage compartment and acknowledgment of underdetected community transmission. To explore this, we expand the standard SEIHFR model to include an isolation-leakage compartment *L* and an undetected infectious compartment *U*, creating the new detection-isolation-leakage SEUIHLF3R model. We fit this model to surveillance data from Ituri Province during the 17th DRC Ebola epidemic using Bayesian methods. The validated model is then used for scenario analysis to estimate the number of cases and deaths averted by interventions, peak isolation needs, and transmission due to leakage. We also analyze how improved case detection and contact follow-up could impact these outcomes.

## Methods and Materials

### Model and Data Sources

The primary data source for our study is the official Situation Reports issued by the Centre d’Opérations d’Urgence de Santé Publique (COUSP) for the period from 14 May to 17 June 2026 (31 situation reports, N*^○^*001-N*^○^*034).^11^ As of 17 June 2026, there were 896 total confirmed cases, 232 confirmed deaths, 78 discharged survivors, and 383 patients in isolation, giving a case fatality ratio of 25.9% among confirmed cases; the reported contact-follow-up rate ranged from 28% to 71% over the series. As of 30 May 2026 (N*^○^*016) the provincial case distribution was 264 confirmed cases in Ituri, 15 in Nord-Kivu, and 3 in Sud-Kivu; we focus on Ituri Province as a self-contained epicenter that accounted for 93.6% of all confirmed cases in eastern DRC, with a laboratory positivity rate of 36.5% (19 of 52 samples) on that date.

The Ebola outbreak had its first suspected case on 24 April 2026. That case was confirmed as a BDBV infection on 15 May 2026. However, the DRC and the WHO announced a Public Health Emergency of International Concern on 17 May 2026. Ebola outbreaks pose many challenges, including refusal of post-mortem testing, incomplete contact tracing, medication shortages, and deficits in isolation capacity. The SitRep report for Ituri province^11^ includes the number of contacts of confirmed cases, the number of contacts successfully followed up, the number of suspected cases tested, the number of hospitalized cases, and the number of escapees.

Summary statistics for the provincial reporting series are shown in Table S1. Over the 14 May-17 June window, cumulative confirmed cases rose from 8 to 896 and confirmed deaths from 4 to 232, with new confirmed cases per inter-report window averaging 30.6 (maximum 107) and new confirmed deaths averaging 7.9 (maximum 42). Escapees were reported on 24 of the reporting days, totaling 175, with a single-day maximum of 23 (on 23 May). The national isolation census, available from 1 June onward, ranged from 173 to 383 patients.

### Epidemiological Model

#### Standard SEIHFR Model

The susceptible-exposed-infectious-hospitalized/isolated-funeral-removed model is the standard model for Ebola. Susceptible individuals *S* become exposed *E* upon infectious contact but are not yet infectious. Upon leaving the exposed compartment, a proportion *p_d_*(*t*) of infections are detected and enter the detected pre-isolation compartment *I*. The remaining proportion, 1 *− p_d_*(*t*), is not detected and enters the undetected community infectious compartment, *U* . Detected infectious individuals in *I* are subsequently isolated or hospitalized in *H*. Infectious individuals in *U* may recover or die in the community. A proportion 1 *− ϕ_D_* of those who die outside a treatment center enter an unsafe funeral compartment *F_U_* . Those who enter *F_U_* receive traditional burial practices, which heighten transmission to their contacts. The remaining *ϕ_D_* of those who die in the community are detected and enter *F_U,D_*. Infectious people who die in isolation centers or hospitals enter a safe funeral compartment *F_H_* . Both *F_U,D_* and *F_H_* receive sanitized burial that does not contribute to transmission. The set of individuals *R* includes only recovered people.

### Isolation-Leakage Extension

In addition to the standard pathway from *I* to *H*, we introduce a *leakage* compartment *L* for individuals who leave isolation *H* at rate *ρ* before fully recovering and then re-enter the community. Individuals in *L* remain infectious in the community at transmission rate *β_L_*. Escapees contribute to community transmission and recover or die at rates *γ_L_* and *µ_L_*, respectively. Deceased escapees enter the unsafe funeral compartment *F_U_* with probability 1 *− ϕ_D_* or the safe funeral compartment *F_U,D_* with probability *ϕ_D_*. This yields the SEUIHLF3R model illustrated in Figure 1.

**Figure 1:**
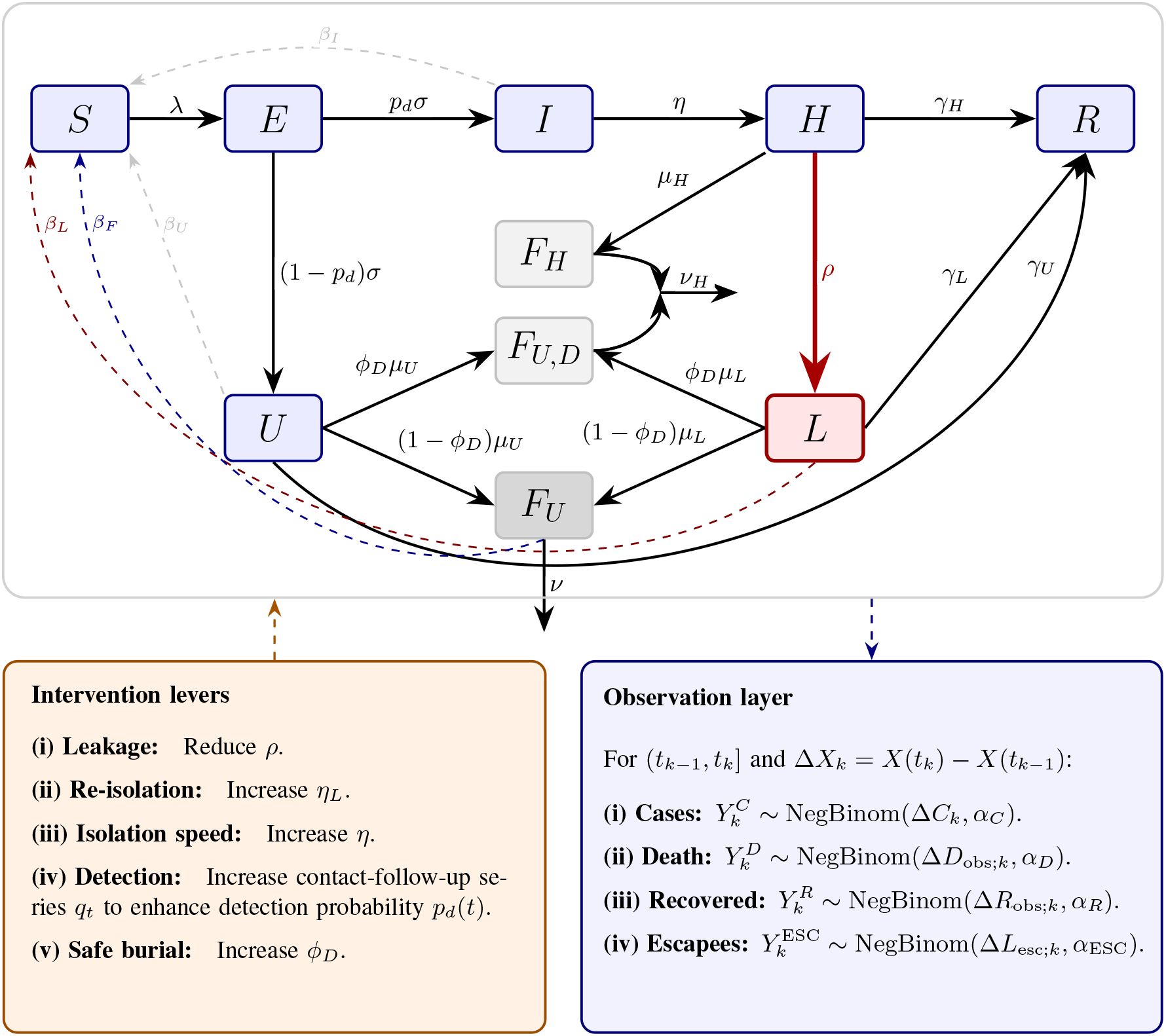
Detection-isolation-leakage SEUIHLF3R model with observation and intervention overlays. Solid arrows represent disease-progression and removal flows, while dashed arrows directed toward the susceptible class represent contributions to the community force of infection. A fraction *ρ* of isolated patients in *H* leaks into the community compartment *L*. The observation layer maps latent transition flows to interval-specific counts of confirmed cases, confirmed deaths, treatment-center discharges, and isolation escape events using negative-binomial observation models. The intervention panel summarizes time-varying detection inputs and counterfactual levers acting on isolation, leakage, burial, and transmission.

The total force of infection is given by

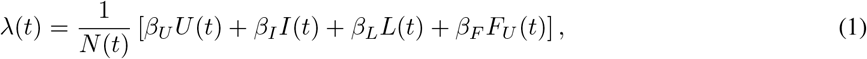

Time variation in transmission enters through a common effective reproduction number *R*_eff_ (*t*) = *R*_0_ *e^−δt^* (a smooth proxy for behavioral change and intervention scale-up; see^12^), so that *β_U_* (*t*) = *R*_eff_ (*t*)*/M* (0) and, by the baseline assumption *β_I_* = *β_L_* = *β_U_* and *β_F_* = *m_F_ β_U_*, all community transmission rates decay together at rate *δ*; *δ* = 0 recovers the constant-*R*_0_ model. Here, *β_U_* represents transmission from undetected infectious individuals in the community, while *β_I_* indicates transmission from detected but not yet isolated infectious persons. The parameter *β_L_*, which is assumed equal to *β_U_*, accounts for post-leakage community transmission. *β_F_* denotes funeral-related transmission from the unsafe burial of the deceased, which is proportional to *β_U_* . Hospitalized patients in *H* do not influence *λ*(*t*), as their isolation is presumed effective. Upon death, hospitalized patients are buried safely and move into *F_H_* ; similarly, a proportion *ϕ_D_* of infected individuals who die in the community are detected and transition into *F_U,D_*, where they are buried safely. Neither *F_H_* nor *F_U,D_* contributes to transmission. The total population size at time *t* is given by *N* (*t*) = *S*(*t*) + *E*(*t*) + *U* (*t*) + *I*(*t*) + *H*(*t*) + *L*(*t*) + *F_U_* (*t*) + *F_U,D_*(*t*) + *F_H_* (*t*) + *R*(*t*).

### SEUIHLF3R Model with Under-detection

The baseline model for BDBV is defined as

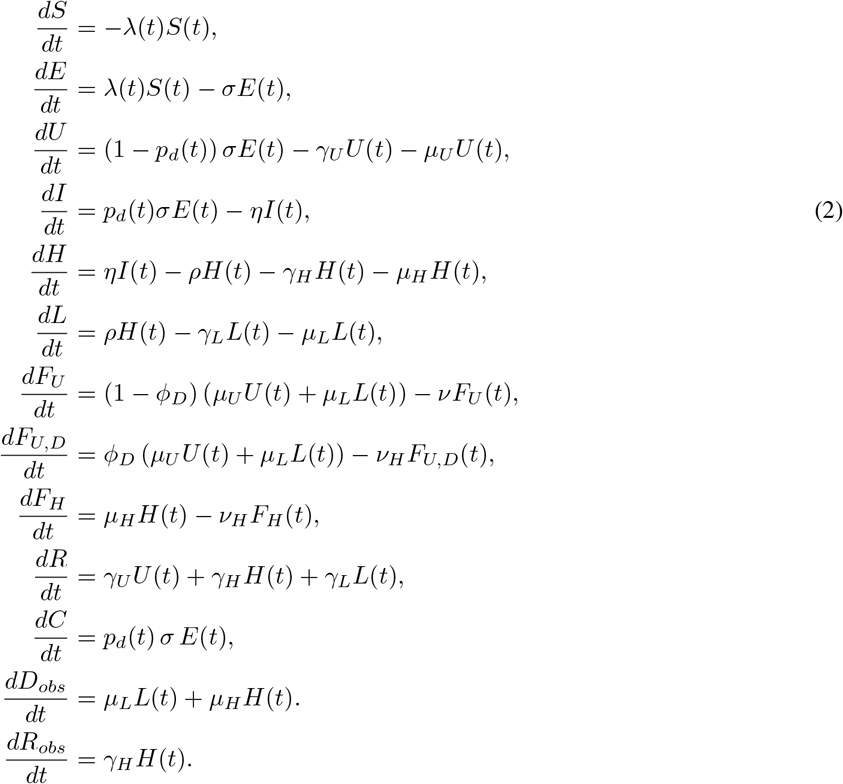

In this model, *F_U_* (*t*) represents the number of undetected infectious people who died outside treatment centers and await unsafe (traditional) burial. They contribute to the community force of infection at rate *β_F_* and undergo unsanitized burial at rate *ν*. The number *F_H_* (*t*) is the count of individuals who died while hospitalized and received sanitized burial at rate *ν_H_* . Similarly, *F_U,D_*(*t*) represents deceased individuals who died in the community but were detected at death and received sanitized burial at the same rate *ν_H_* . Safely buried individuals do not contribute to transmission, and the exact value of *ν_H_* does not affect the outbreak dynamics. The leakage rate *ρ* governs how quickly hospitalized patients exit isolation. The confirmed-death accumulator *D_obs_* counts deaths only among laboratory-confirmed cases, matching the SitRep definition (*décès parmi les confirmés*), isolated patients (*H*) are confirmed at detection, and escapees (*L*) were confirmed before absconding, so both contribute to *D_obs_*. Undetected community deaths (from *U*) are never laboratory-confirmed; they are recorded as probable deaths and therefore do not enter *D_obs_*. Consequently, *D_obs_* is decoupled from the burial routing as *ϕ_D_* governs only whether a community death receives a safe (non-transmitting, *F_U,D_*) versus an unsafe (transmitting, *F_U_*) burial, and does not enter the observed-death series. The discharge accumulator *R_obs_* collects treatment-center recoveries only (*γ_H_ H*); community self-recoveries in *U* and escapees in *L* are never counted as discharges.

### Escapees sojourn times

Patients who leave isolation before clinical resolution enter the leaked compartment *L*. We impose the biological consistency constraint that the total expected time from hospitalization to disease resolution is the same regardless of whether the patient remains in *H* or escapes to *L*,

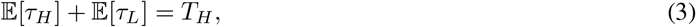

where *T_H_* = 10.8 days is the mean hospitalization duration. Under exponential sojourn times, E[*τ_H_*] = (*γ_H_* + *µ_H_* + *ρ*)*^−^*^1^, so the required total exit rate from *L* is

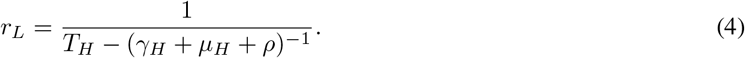

The recovery-death split within *L* mirrors that of *H*, since an escaped patient’s prognosis is conditioned on hospital stage disease rather than community-stage disease,

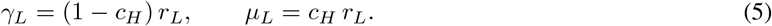

Note that *r_L_* increases with *ρ* as faster leakage implies a shorter mean stay in *H* and therefore a longer remaining disease course in *L*. Intuitively, an escapee has already undergone part of their infectious period in isolation. As a result, the time they remain infectious within the community is reduced, and the earlier they escape, the larger the fraction of their disease course that unfolds outside.

In the base model, we set 1*/γ_L_* + 1*/ρ* = 1*/γ_H_* and 1*/µ_L_* + 1*/ρ* = 1*/µ_H_* such that *ρ >* max(*γ_H_, µ_H_*). See Figure 1 for a schematic illustration of the model.

For more information about the model assumptions, see the Supplementary Material S1.2.

### Basic and Detection-Conditioned Reproduction Numbers

Let the basic reproduction number be *R*_0_, and the detection-conditioned reproduction number be *R*(*p_d_*). The basic reproduction number is defined in the absence of detection and isolation (*p_d_* = 0); that is, *R*_0_ = *R*(0). It represents the reference community transmissibility of BDBV in a fully susceptible population before detection, isolation, and safe burial interventions are implemented. In contrast, *R*(*p_d_*) describes transmission under detection probability *p_d_* and decomposes transmission into undetected community, detected pre-isolation, leakage, and funeral components. This distinction is important because previous Ebola modeling studies often estimated reproduction numbers from outbreaks in which detection, isolation, contact tracing, and burial interventions were already in effect.^27,36,16^ For the detection-isolation-leakage SEUIHLF3R model, the next-generation matrix approach^33,9^ gives

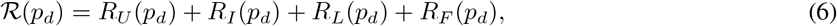

where

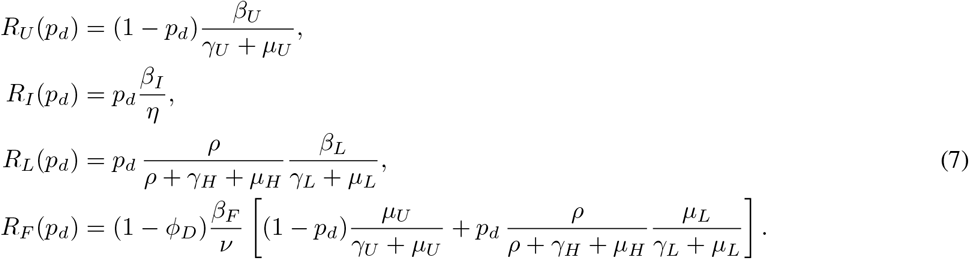

Setting *ρ* = 0 removes the leakage contribution (*R_L_* = 0); setting *p_d_* = 1 omits the undetected community pathway *U*, yielding the detected-only reproduction number. In the baseline parameterization, we assume *β_I_* = *β_U_*, *β_L_* = *β_U_*, and *β_F_* = *m_F_ β_U_* with *m_F_ ≥* 1. Therefore, the detection-conditioned reproduction number factors as *R*(*p_d_*) = *β_U_ M* (*p_d_*), where *M* (*p_d_*) = *M_U_* (*p_d_*) + *M_I_* (*p_d_*) + *M_L_*(*p_d_*) + *M_F_* (*p_d_*), and *M_T_* (*p_d_*) = *R_T_* (*p_d_*)*/β_U_* for *T ∈ {U, I, L, F}*.

### Observation Model

Reported Ebola cases are often delayed, underreported, and frequently reclassified from suspected to confirmed once laboratory results are received. Under the extended model, the detected incidence is 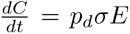; undetected cases in *U* are not observed and contribute only to true but unreported infections and to *F_U_* and *F_U,D_* upon death. Let *Y ^C^*(*t*) denote the observed daily number of reported cases, and *Y ^D^*(*t*) the daily number of reported deaths at time *t*. We use negative binomial count likelihoods to model these observations, accounting for overdispersion. That is,

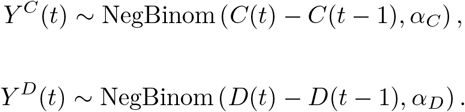

The dispersion parameters *α_C_* and *α_D_* account for superspreading, reporting heterogeneity, delayed confirmation, and surveillance noise. The likelihood is placed on increments between successive situation reports (29 windows over 14 May-17 June), obtained by differencing the simulated cumulative solution at the two report days; because reports are unequally spaced, this integrates the detection flow *p_d_*(*t*)*σE*(*t*) over the actual window length without any fixed-Δ*t* assumption. Windows in which a reported cumulative count decreases (case reclassification between reports) are dropped from the likelihood rather than clipped to zero.

Cumulative treatment-center discharges (*guéris*) are reported alongside cases and deaths. Because deaths and recoveries are the two competing exits from isolation *H*, observing only the death arm leaves their ratio weakly identified; we therefore add a discharge likelihood on the observable-recovery accumulator *R_obs_*,

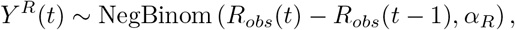

which pins the in-isolation recovery/death split.

The observed escapee count *Y ^ESC^*(*t*) is modeled as

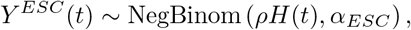

where *H*(*t*) is the reported provincial isolation census (*patients en isolement/hospitalisés*). This links the leakage rate *ρ* to the reported daily escapee counts and allows overdispersion across days via *α_ESC_*. The stream contributes to the 15 reports (1-17 June 2026) where both escapees and the isolation census are recorded, 97 escapees in total, and estimates *ρ* independently of the incidence, death, and discharge data streams.

The situation reports provide both reported and suspected cases, those tested because of symptoms, and contacts listed and followed after exposure to confirmed cases. We therefore decompose the detection probability among newly infectious individuals into two non-exclusive routes, which are symptom-based surveillance/testing and contact-tracing-based detection. Let *q_t_* denote the reported contact-follow-up proportion and let *z_t_* denote a standardized proxy for symptom-based surveillance intensity, constructed from suspected cases, alerts, or testing activity when available. We define

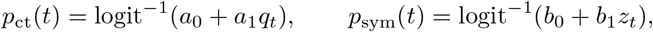

and combine them as

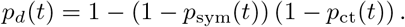

This specification allows the data to estimate time-varying detection while respecting the known natural-history timescale.

### Estimation and Simulation

We solve the system of ordinary differential equations (ODEs) using a fourth-order Runge-Kutta method. We use Bayesian inference to estimate a restricted set of parameters to ensure identifiability. In particular, we estimate the leakage rate *ρ*, primarily identified from the escapee count in the Bunia facility data; the total transmission intensity via the basic reproduction number *R*_0_; the detection parameter *p_d_*(*t*); and the overdispersion parameters *α_C_*, *α_D_*, and *α_ESC_*. In the baseline analysis, we fix the natural-history and funeral-process parameters, *T_U_*, *c_U_*, *γ_U_*, *µ_U_*, *ν*, *ν_H_*, and *m_F_*, and vary them in scenario and sensitivity analyses. We use externally specified community infectious duration *T_U_* and community fatality probability *c_U_* to reduce structural non-identifiability in the undetected and funeral-transmission pathways. Specifically, we set *γ_U_* = (1 *− c_U_*)*/T_U_* and *µ_U_* = *c_U_ /T_U_* . Similarly, we fix the unsafe-burial removal rate *ν* and the relative funeral transmissibility *m_F_* = *β_F_ /β_U_* based on the literature. These restrictions are necessary because the reported case and death time series cannot separately identify the undetected burden, the community fatality probability, the death-detection probability, and the funeral infectiousness.

Parameters include the incubation rate *σ*, infectious-period rates, the time from detection to isolation, the safe-burial rate, and the disease-outcome rates. These are set using literature values from the 2007 BDBV Uganda outbreak^32,17,37,29^ and from a systematic review of Ebola modeling parameters.^24^ For more information on parameter assignments and prior distributions, see the Supplementary Material S1.3.

Historically, BDBV-specific estimates of *R*_0_ have been scarce.^24^ However, a CDC scenario-projection analysis of the current 2026 Bundibugyo virus disease outbreak reported a calibrated median *R*_0_ = 2.51 with an interquartile range of 2.27*−*2.82, based on 50 deaths by 24 May 2026.^21^ Therefore, we use this estimate to inform the prior range for *R*_0_, while recognizing that the CDC branching-process model differs structurally from our compartmental detection-isolation-leakage model. We use historical estimates from Zaire ebolavirus outbreaks in the DRC and other countries for sensitivity analyses, as they reflect different species, intervention histories, and surveillance conditions.^8,27,36,23^ Accordingly, we use a prior for the basic reproduction number, given by

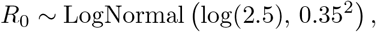

which places most prior mass near the CDC’s current outbreak interquartile range. The affected-population scale is fixed at *N* (0) *≈* 2,000,000, approximating the Ituri health zones with active transmission.

We implemented the model in Python using PyMC,^30^ with ordinary differential equation (ODE) solutions provided by NumPy and SciPy.^35^ We use Bayesian methods to assess whether leakage and funeral components are supported by the data beyond what is expected from the priors alone. In particular, we use posterior predictive checks and the Watanabe-Akaike information criterion (WAIC) to compare the SEUIHLF3R model to its nested SEIHFR model with no leakage (*ρ* = 0) and to the simpler model with no funeral compartments.

For more information about the model’s parameters, see the Supplementary Material S1.3.

The codes and datasets used and/or analyzed in this paper are available through https://github.com/ tamfatkh/ebola.

### Model, Intervention and Scenario Analyses

#### Reproduction-number decomposition and control boundaries

Using the next-generation decomposition in (6)-(7), we evaluated the detection-conditioned reproduction number *R*(*p_d_*) = *R_U_* + *R_I_* + *R_L_* + *R_F_* . We estimated the posterior contributions of each component at different detection levels of *p_d_*. We then examined its sensitivity to detection *p_d_*, leakage *ρ*, and death ascertainment *ϕ_D_*, each varied about its posterior median.

To characterize joint control requirements, we mapped the *R* = 1 contour in the detection-isolation plane (*p_d_, η*) at median *ϕ_D_*, for a range of leakage rates. Because faster isolation removes only the pre-isolation contribution *R_I_* = *p_d_β_U_ /η*, whereas the undetected (*R_U_*) and leakage (*R_L_*) contributions are independent of *η*, control is unattainable below a detection floor 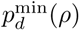, defined as the smallest *p_d_* for which *R <* 1 in the limit *η → ∞*; above a threshold leakage rate no (*p_d_, η*) combination attains *R <* 1.

### Counterfactual framework

We evaluated control measures using forward counterfactual simulation based on the fitted posterior and without re-estimation. For each posterior draw *s* = 1*, . . ., S* of the parameter vector

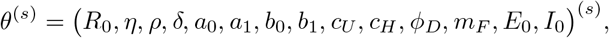

we simulated the SEUIHLF3R system (2). Under each scenario, one or more parameters were replaced by their counterfactual values while all remaining components of *θ*^(*s*)^ were held at their posterior values. The counterfactual simulator reproduces the fitted reproduction multiplier *M* (0) and derived progression, mortality, and funeral rates, as well as the burial routing and the escapee sojourn-time construction. It also reproduces the exponential trend *R*_eff_ (*t*) = *R*_0_*e^−δt^*. The two-route detection probability *p_d_*(*t*) is driven by the same daily contact follow-up *q_t_* and symptom-surveillance proxy *z_t_* used in estimation.

For each scenario, we recorded cumulative actual infections (*σE dt*), cumulative reported cases (*p_d_σE dt*), cumulative reported deaths, the reported hospitalization *H*(*t*) and its peak (peak isolation demand), and the leakage-attributable share of transmission. Intervention impact was measured as actual infections and deaths averted relative to the fitted-*ρ* baseline, along with the corresponding percentage reduction. Propagating each scenario over the full set of draws preserves the joint posterior dependence among parameters. Every projected trajectory and derived summary computed across draws is reported using the posterior median and a 95% highest-density interval (HDI).

### Intervention levers

We considered five intervention levers, each acting on a distinct mechanism of the Ebola spreading system.

i. *Leakage* was explored by changing the escape rate *ρ* by a multiple 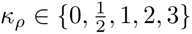. It was also explored via a time-varying step *ρ*(*t*) that switches to zero after week 1, 2, 3, or 4 from its posterior median.
ii. *Re-isolation* of escaped patients was introduced through a return-to-isolation rate *η_L_*. It moves individuals from the community leakage compartment *L* back into the isolation compartment *H*. It changes the following two equations

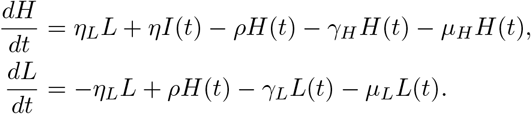

Re-isolation scenarios applied constant return-to-care rates *η_L_ ∈ {*0, 0.05, 0.10, 0.20*}* day*^−^*^1^. The scenarios correspond to mean recapture times of *∞,* 20, 10, 5 days.

(iii) *Isolation speed* was increased by scaling the detected-to-isolation rate by a multiple *κ_η_ ∈ {*1, 1.5, 2, 3*}*. That decreases the pre-isolation community-transmission period and accelerates hospitalization.
(iv) *Detection* was strengthened by scaling the reported contact-follow-up series, *κq_t_* with a multiple *κ ∈* [1, 3], which increases the probability of detection by contact tracing *p*_ct_(*t*). It in turn increases the combined detection probability *p_d_*(*t*) = 1 *−* (1 *− p*_sym_(*t*)) (1 *− p*_ct_(*t*)). An idealized benchmark *p_d_* was alternatively fixed at a constant.
(v) *Safe burial* was improved by raising the (ascertainment) post-mortem detection/safe-burial probability *ϕ_D_*. It re-allocates community deaths from the transmitting unsafe-funeral compartment *F_U_* to the sanitized compartment *F_U,D_*. The examined values of *ϕ_D_* are above its fitted value.

In the detection scenarios analysis in (iv) and ascertainment scenarios analysis in (v), we asked how much of the leakage-attributable burden could be recovered, benchmarked against the *no-leakage target*, defined as the cumulative actual infection burden under *ρ* = 0.

There are two important changes under those scenarios, in particular re-isolations in (ii) and safe burial in (v).

First, when re-isolation is active, a re-isolated escapee can leak again, so the isolation and community-leakage compartments *H* and *L* form a cycle, and the leakage pathway acquires a geometric correction. Writing

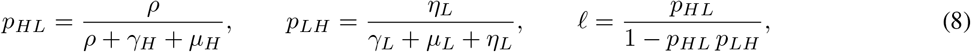

where *p_HL_* is the per-*H*-visit probability of leaking, *p_LH_* is the per-*L*-visit probability of return to hospital, and *ℓ* is the expected number of community-leakage episodes (*L*-visits) per detected case. Thus, the leakage and funeral contributions to the detection-conditioned reproduction number in (7) become

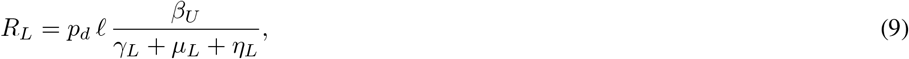

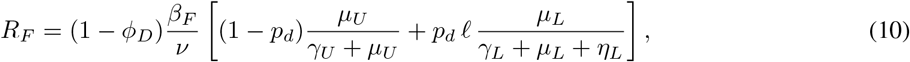

with each *L*-visit transmitting for a mean time (*γ_L_* + *µ_L_* + *η_L_*)*^−^*^1^ and ending in death with probability *µ_L_/*(*γ_L_* + *µ_L_* + *η_L_*). Setting *η_L_* = 0 gives *p_LH_* = 0 and *ℓ* = *ρ/*(*ρ* + *γ_H_* + *µ_H_*), recovering the baseline expressions

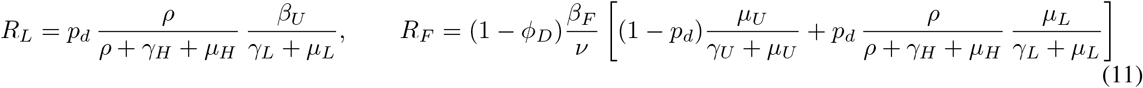

of (7). The undetected and pre-isolation components *R_U_* and *R_I_* do not involve the leakage cycle and are unchanged.

Here, *η_L_* acts as an additional competing exit from *L* that leaves the disease-resolution rates *γ_L_, µ_L_* unchanged. Therefore, the geometric cycling in (8) thus assumes memoryless re-entry to *H*, which relaxes the single-pass sojourn constraint E[*τ_H_*] + E[*τ_L_*] = *T_H_* once re-isolation is active.

Second, the burial lever requires care because the community transmission rate is tied to *R*_0_ through *β_U_* = *R*_0_*/M* (0), and *M* (0) contains the funeral term (1 *− ϕ_D_*)*m_F_* (*µ_U_ /*(*γ_U_* + *µ_U_*))*/ν*. For the descriptive reproduction-number decomposition and its uncertainty (below) we retained the estimation convention, in which *β_U_* is re-anchored so that *R*_0_ is held fixed as *ϕ_D_* is varied. For the burial *intervention* scenario, however, we held *β_U_* at its fitted value and let *ϕ_D_* enter only the death-routing terms, so that improved safe-burial coverage lowers the funeral force of infection and reduces *R* as a consequence, rather than being offset by a compensating increase in *β_U_* . Holding *β_U_* (per-contact transmissibility) fixed while burial practice changes, and allowing *R*_0_ to fall as an outcome, is the appropriate convention for an intervention, and the two conventions coincide at the fitted *ϕ_D_*.

## Results

### Model fitting and analysis using Bayesian Methods

We applied a Markov Chain Monte Carlo (MCMC) framework to conduct Bayesian inference and estimate the model’s parameters. Prior predictive checks (Figure S1) demonstrated that the priors produce trajectories for cases, deaths, recoveries, and escapees that match the overall scale of the observed outbreak and exhibit excellent coverage. The MCMC trace plots (Figure S2), together with visual inspection, rank statistics (*r̂* < 1.01), bulk and tail effective sample sizes (*ESS >* 400), support convergence for all inferred parameters (Table S4). The posterior contractions and shifts relative to the priors in Table S3 suggest that the data were informative for these parameters (Figure S3). See detailed posterior summaries and diagnostics in Supplementary Material S1.4. Finally, the posterior median and 95% high-density interval (HDI) in Figure 2 indicate that the model reproduces the empirical patterns in reported cases, deaths, recoveries, and escapees. The posterior median trajectories closely follow the reported series, and the 95% HDIs expand over the observation period.

**Figure 2:**
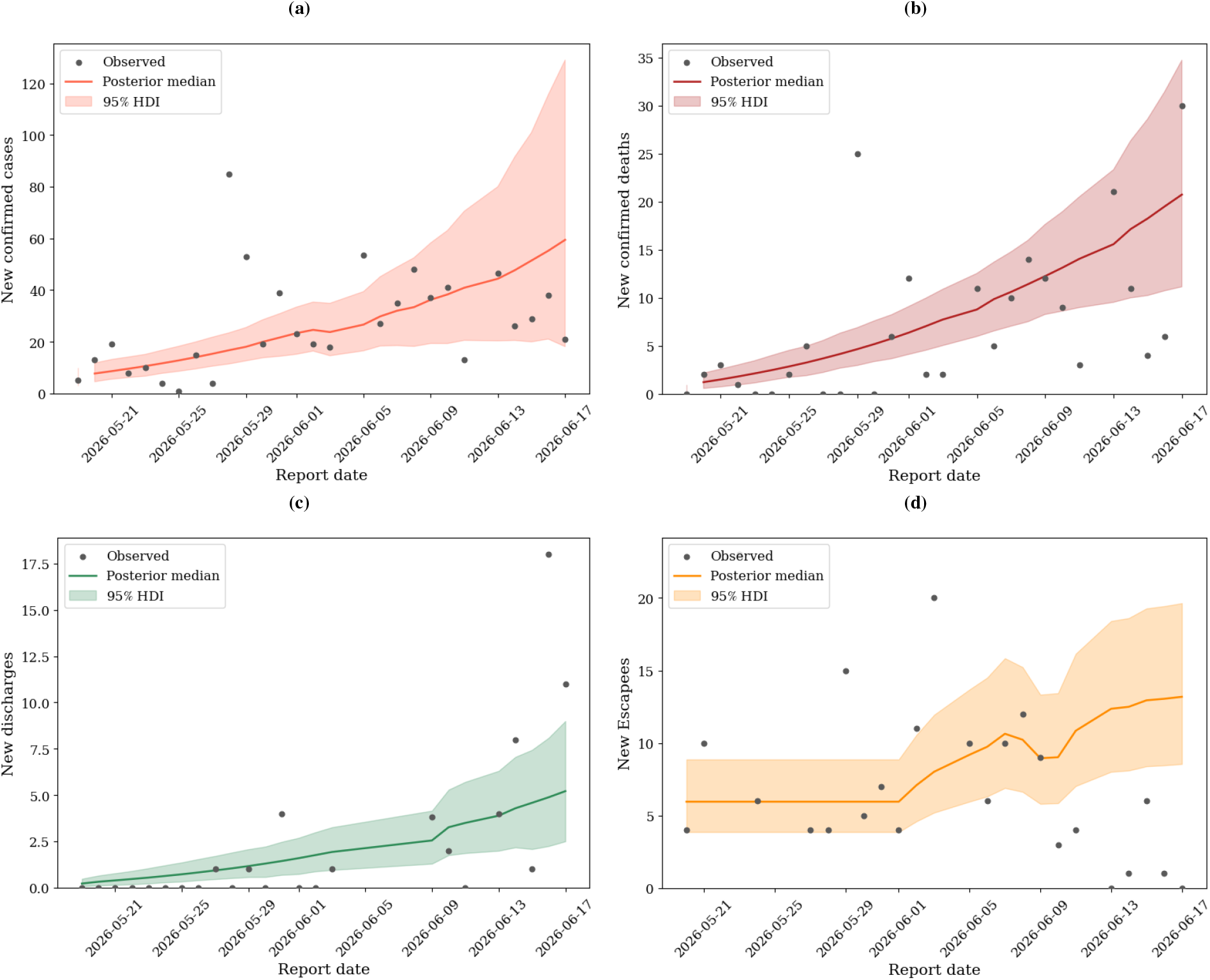
Median and 95% HDI of the posterior predictive versus reported data of (a) the number of Ebola cases, (b) the number of deaths, (c) the number of recoveries, and (d) the number of escapees in Ituri province with parameters estimated using Bayesian methods.

Figure 3 shows the estimated detection probability *p_d_*(*t*) along with its two components, symptom-based surveillance *p*_sym_(*t*) and contact tracing *p*_ct_(*t*). The slope coefficients linking each stream to detection *a*_1_ and *b*_1_ were credibly positive, with 95% HDIs excluding zero. However, the intercepts *a*_0_ and *b*_0_ were not resolved, with their HDIs including zero. The combined probability *p_d_* = 1 *−* (1 *− p*_sym_)(1 *− p*_ct_) is dominated by whichever stream is higher. Seemingly, contact tracing remained consistently high and stable with *p*_ct_ *≈* 0.84, and so detection was driven mainly by the contact tracing stream. Meanwhile, a sharp drop in the symptom stream in early June, with *p*_sym_ falling from about 0.97 to 0.33, translated into only a modest decline in *p_d_*, from about 0.99 to 0.91.

**Figure 3:**
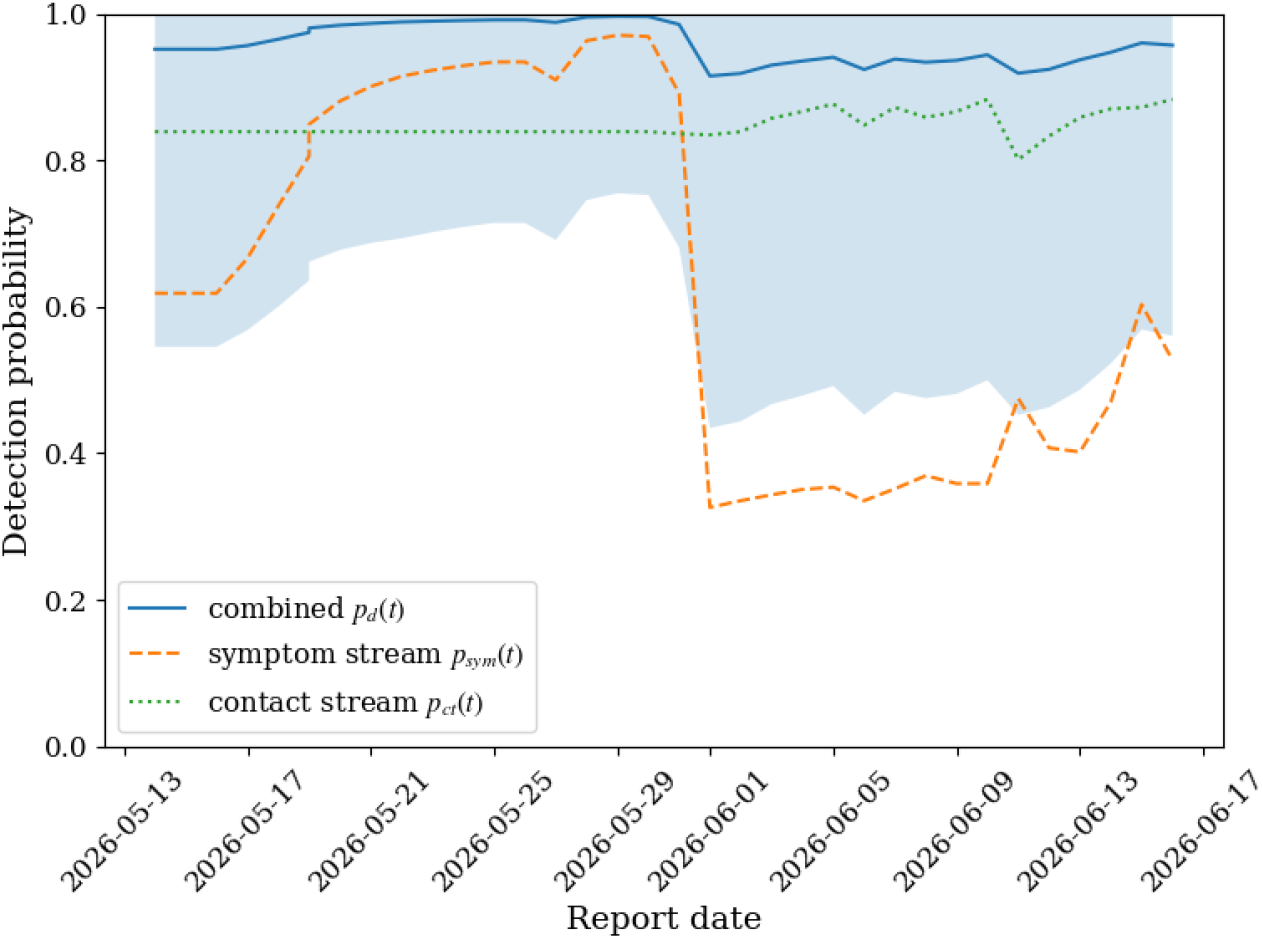
Estimated median detection probability and its 95% HDI.

Figure 4 shows the sensitivity of *R*(*p_d_*) to detection probability *p_d_*, leakage *ρ*, and death ascertainment *ϕ_D_* across their ranges, while holding the other two at their posterior medians. The vertical dotted line in the middle panel of Figure 4 marks the fitted *ρ̂* posterior median value (*ρ* = 0.034). Leakage has by far the strongest effect on *R*, which increases monotonically from 3.2 at *ρ* = 0 to above 5 across its range. It declines only slightly with detection *p_d_* and increases weakly with *ϕ_D_*. The fact that *R* remains above one, with wide posterior bands excluding *R* = 1 throughout all three ranges, reinforces that no single intervention lever can contain the outbreak on its own.

**Figure 4:**
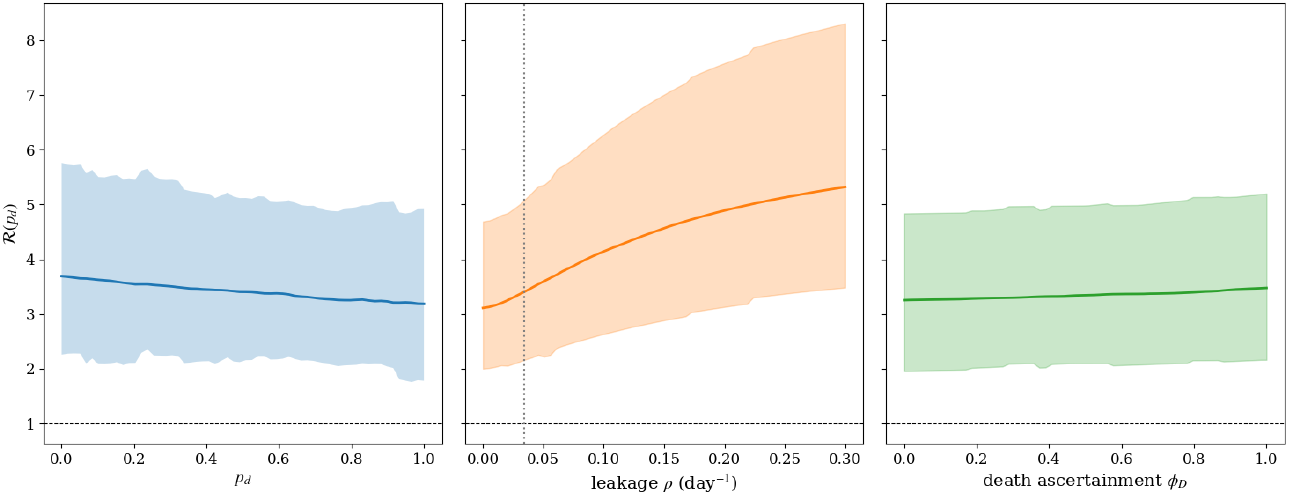
Sensitivity of the median *R*(*p_d_*) to *p_d_*, *ρ*, *ϕ_D_* (one at a time) and the corresponding 95% HDI .

Figure 5 (a) breaks down the detection-conditioned reproduction number *R*(*p_d_*) into its undetected *R_U_*, pre-isolation *R_I_*, leakage *R_L_*, and funeral *R_F_* components. At low detection probability *p_d_*, the undetected community pathway dominates. But as *p_d_* increases, this contribution is gradually shifted to the pre-isolation pathway *R_I_* . Under our assumption, detected and undetected cases transmit at the same rate until isolation, increase case detection mainly shifts infections between the two compartments (*I, U*) with similar transmissibility. Raising *p_d_* only reduces *R* from about 3.7 at *p_d_* = 0 to 3.2 at *p_d_* = 1. Thus, detection alone cannot achieve control; see also the first panel in Figure 4. The contributions of leakage and funeral components are relatively small, with leakage contributing more than funeral transmission.

**Figure 5:**
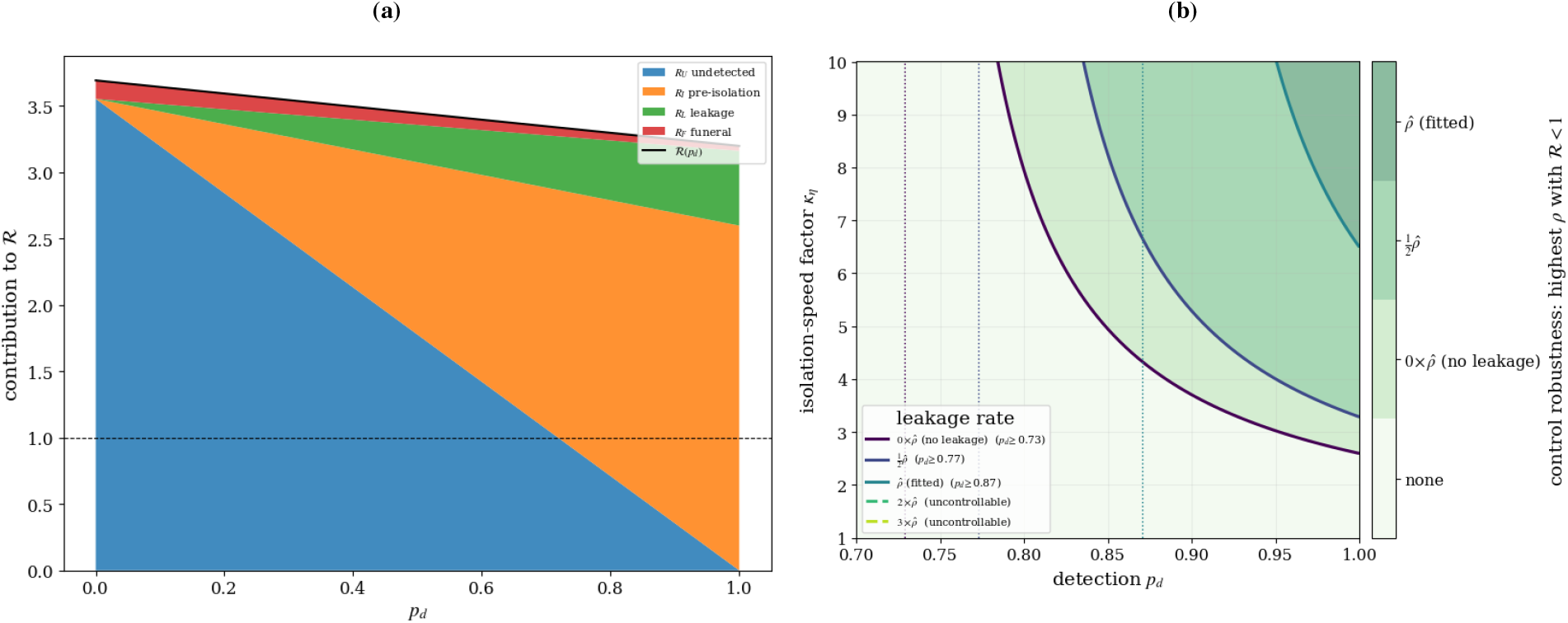
(a) Decomposition of *R*(*p_d_*) into *R_U_, R_I_, R_L_, R_F_* at *ρ* = 0.034. (b) equi-lines of *R*(*p_d_, κ_η_*) = 1 at different leakage rates *ρ* showing the least possible detection probability at which the Ebola outbreak will be curbed. The term "none" on the color bar of panel (b) means the disease is not under control, while in the other regions it is controlled at the respective values of *ρ*.

Figure 5 (b) shows the *R* = 1 control boundary in a plane defined by the detection probability *p_d_* and the isolation-speed factor *κ_η_* = *η/η̂*. The detection-isolation plane is built using posterior-median parameters, with leakage shown as multiples of the fitted rate *ρ̂*. For each leakage level, control with *R <* 1 is achieved only above and to the right of the corresponding contour. This means containment depends on both sufficient case detection and sufficiently rapid isolation. The shaded gradient summarizes the robustness of control to leakage rates. The shading depth at any point indicates the highest leakage rate for which that combination of *p_d_* and *κ_η_* still gives *R <* 1. As leakage increases from 0 to 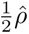, the boundary shifts toward higher *p_d_* and higher *κ_η_*. Greater escape from isolation must be offset by better detection and faster isolation to reach the same threshold. In other words, leakage makes control monotonically harder.

Furthermore, in Figure 5 (b), each contour approaches a vertical asymptote, the *detection floor*, which represents the minimum detection probability below which no isolation rate can reduce *R* below one. This floor arises because increasing the isolation rate only decreases the pre-isolation component *R_I_*, whereas the undetected-community *R_U_* and leakage *R_L_* components are independent of *κ_η_*. When detection becomes too low, undetected infections and leakages are sufficient to maintain transmission regardless of how rapidly detected cases are isolated; see also panel (a). The height of this floor grows steeply with leakage, from *p_d_ ≥* 0.73 at *ρ* = 0 to *p_d_ ≥* 0.77 at 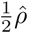 and *p_d_ ≥* 0.87 at the fitted leakage rate *ρ̂*. For leakage levels above the fitted rate, there is no viable solution in this parameter plane. Specifically, at both 2*ρ̂* and 3*ρ̂*, no pairing of (*p_d_, κ_η_*) yields *R <* 1. Thus, once leakage is roughly doubled relative to its fitted value, improvements in detection and isolation speed alone are no longer sufficient. In such a case, successful containment additionally requires reducing leakage, either by reducing *ρ* or by swiftly re-isolating escaped patients *η_L_*.

We used the fitted model to run counterfactual scenarios, reintegrating the posterior over a range of leakage, re-isolation, isolation speed, detection, and safe-burial settings while keeping all other parameters at their posterior values.

Figure 6 (a) scales the leakage rate *ρ* by multiples of the estimated *ρ̂*, and Table 1 lists the associated end-of-window burdens (total counts). Across all intervention measures, leakage substantially amplifies transmission. The trajectories remain monotonically ordered with respect to *ρ*, and the 95% HDIs expand as leakage increases, reflecting the compounded uncertainty arising from accelerated community transmission. Removing leakage altogether with *ρ* = 0 lowers cumulative actual (reported and unreported) infections from 1,120 at the fitted rate to 764. In contrast, doubling and tripling leakage increase cumulative infections to 1,478 and 1,800, respectively. Reported cases are the most sensitive, climbing from 677 with no leakage to 1,575 at 3*ρ̂*. That might be due to the fact that escapees who remain infectious in the community initiate new detected counts. Reported deaths, by comparison, grow more moderately from 240 to 350 across the same range.

**Figure 6:**
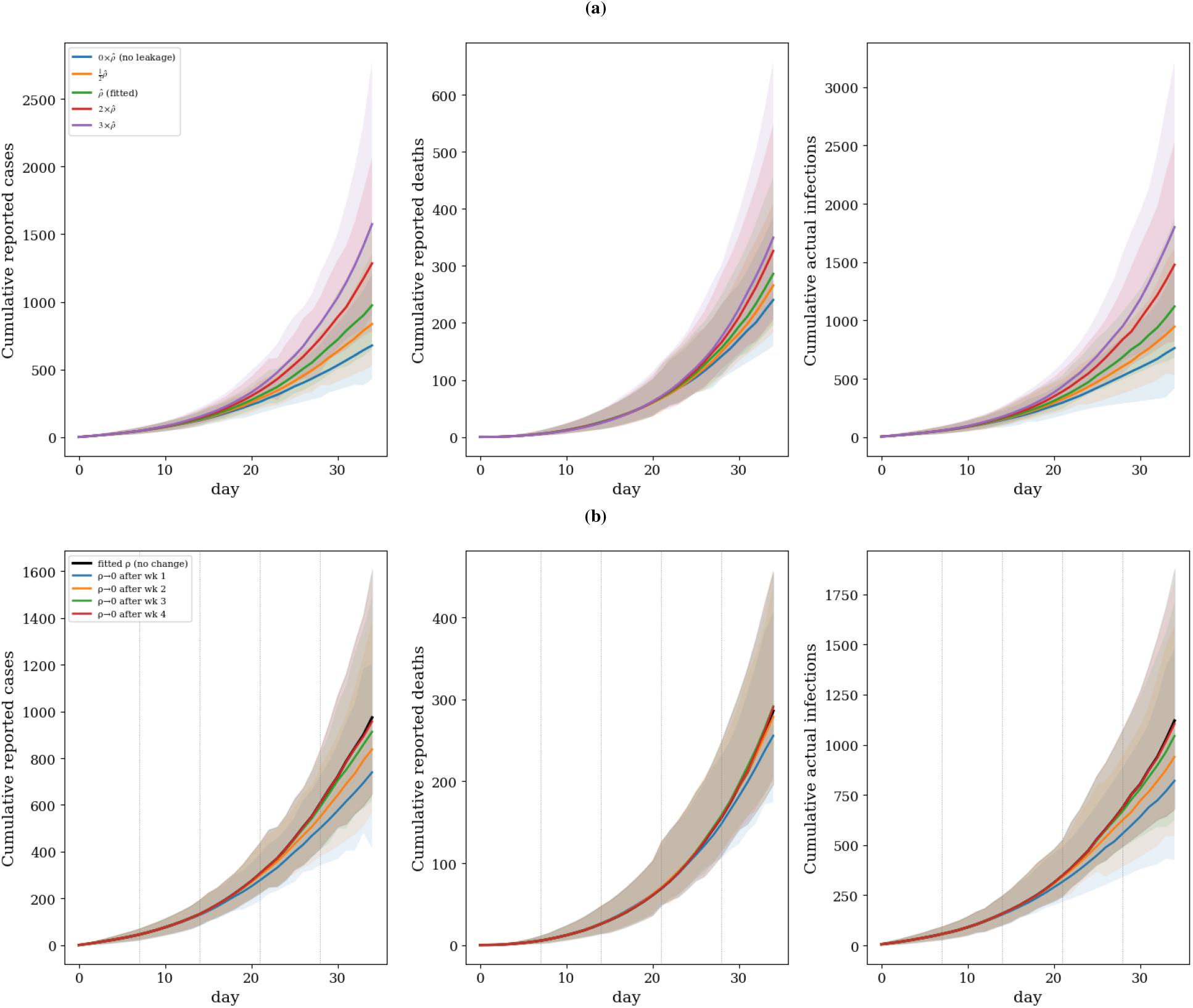
Cumulative reported cases, reported deaths, and actual infections under (a) counterfactual leakage rates *ρ*, relative to the fitted *ρ*, and (b) drop of leakage rates *ρ* to 0, preceded by the fitted *ρ*, with 95% HDI. Actual infections is the sum of both reported and unreported cases.

**Table 1:** Reported cases, reported deaths, and actual infections under different leakage scenarios. Actual infections is the sum of both reported and unreported cases.

| Scenario ($\kappa_\rho \times \hat{\rho}$) | Reported Cases | Reported Deaths | Actual Infections |
| --- | --- | --- | --- |
| $0 \times \hat{\rho}$ (no leakage) | 677 | 240 | 764 |
| $\frac{1}{2} \times \hat{\rho}$ | 850 | 265 | 937 |
| $\hat{\rho}$ (fitted) | 974 | 286 | 1,120 |
| $2 \times \hat{\rho}$ | 1,284 | 326 | 1,478 |
| $3 \times \hat{\rho}$ | 1,575 | 350 | 1,800 |

Because leakage accumulates over time, the benefit of suppressing it depends heavily on how early the intervention begins. Figure 6 (b) and Table 2 show the effect of setting *ρ* to zero after different intervention weeks. Eliminating leakage after week 1 prevents 300 actual infections, or about 26.7% relative to the fitted-*ρ* trajectory. This benefit, however, diminishes quickly with delay, with 180 infections averted (16.1%) after week 2, 76 (6.8%) after week 3, and only 16 (1.5%) after week 4. In Figure 6 (b), the corresponding trajectories deviate noticeably from the unchanged fitted-*ρ* curve only when the switch occurs early. The trajectories gradually converge to it as the intervention is postponed. The reason is that, by the end of the observation period, most leakage-driven transmission has already occurred. This sharp drop-off highlights that controlling leakage is primarily useful as an early-response strategy.

**Table 2:**
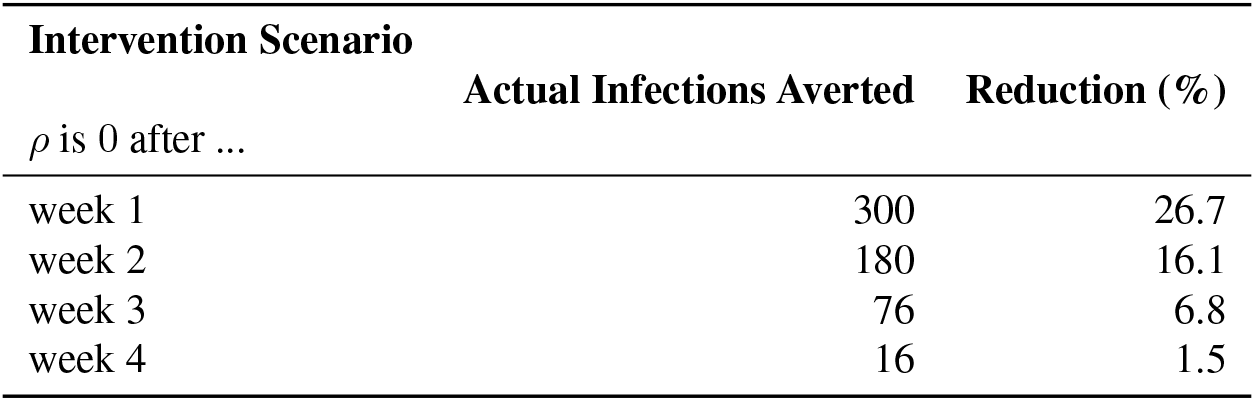
Averted actual infections relative to the fitted-*ρ* scenario, by intervention week. Values are posterior medians.

| Intervention Scenario | Actual Infections Averted | Reduction (%) |
| --- | --- | --- |
| $\rho$ is 0 after ... | | |
| week 1 | 300 | 26.7 |
| week 2 | 180 | 16.1 |
| week 3 | 76 | 6.8 |
| week 4 | 16 | 1.5 |

Figure 7(a) and Table 3 analyze the complementary strategy of reducing onward transmission by re-isolating leaked patients at rate *η_L_*. Here, 1*/η_L_* represents the mean number of days required to recapture an escapee. Larger values of *η_L_* correspond to a quicker return from the community to non-transmitting isolation. Increasing the re-isolation rate produces moderate but growing benefits with 49 actual infections averted (4.4%) at *η_L_* = 0.05 day*^−^*^1^, 99 (8.9%) at 0.10 day*^−^*^1^, and 158 (14.1%) at 0.20 day*^−^*^1^, which corresponds to a mean recapture time of five days. As with eliminating leakage, the timing of implementation is important. Activating *η_L_* = 0.20 day*^−^*^1^ only after week 1 prevents 149 infections (13.3%), but this impact shrinks to 106 (9.4%), 44 (3.9%), and 13 (1.1%) when activation is delayed until weeks 2, 3, and 4, respectively (Table 4). Importantly, continuous re-isolation from the outset yields only a slightly larger effect than initiating it in week 1, with 14.1% versus 13.3% infections averted, since the difference lies only in how leakage is handled in the first week. Moreover, both strategies are less effective than eliminating leakage entirely. This is expected, because re-isolation can only recover a fraction of individuals who have already escaped, rather than avoiding escape events altogether.

**Figure 7:**
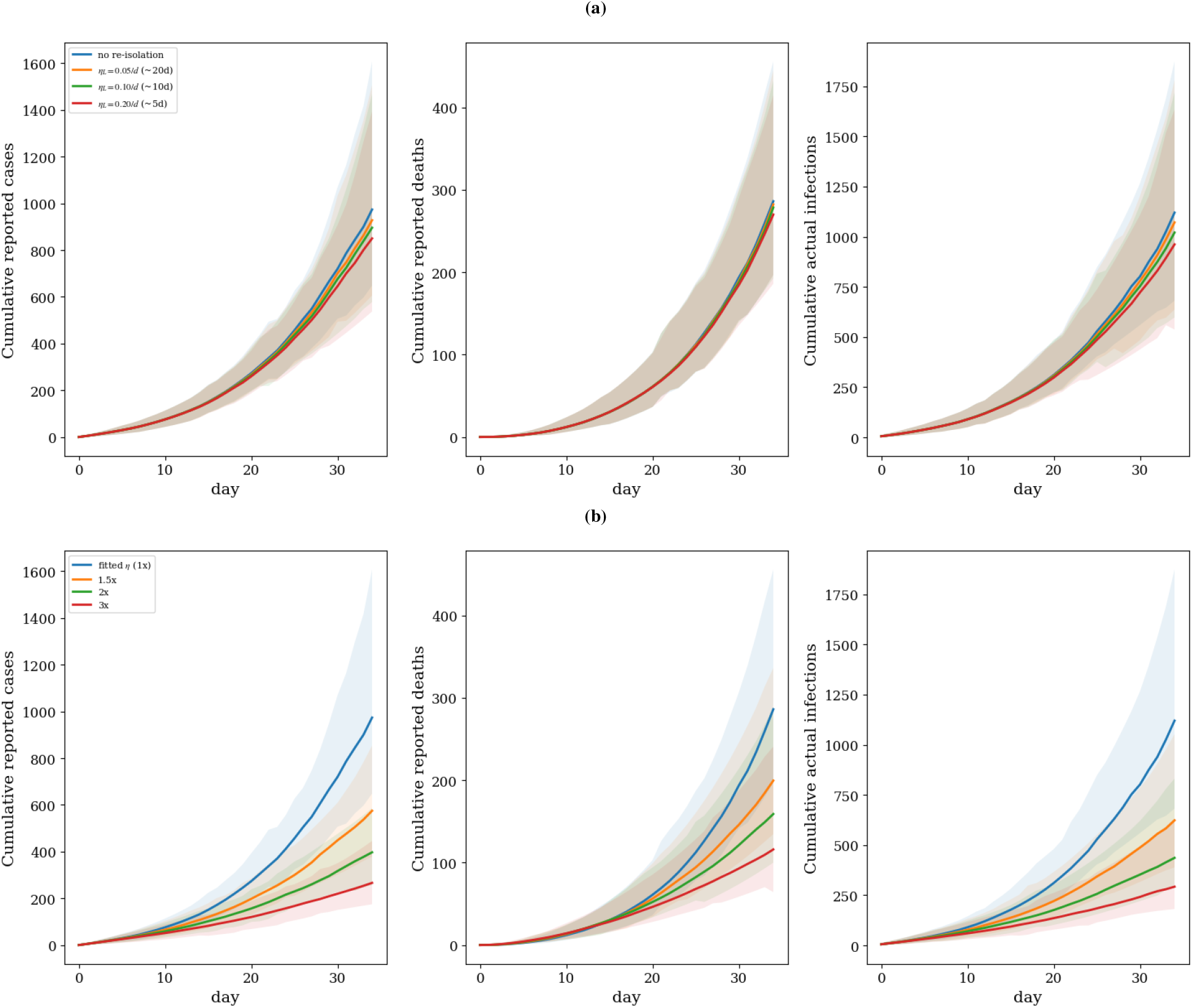
Cumulative reported cases, reported deaths, and actual infections under (a) counterfactual re-isolation rates *η_L_* and (b) counterfactual isolation rates *η*, relative to the fitted *η* with 95% HDI.

**Table 3:** Averted actual infections under constant re-isolation of leaked patients, relative to no re-isolation (*η_L_* = 0). Values are posterior medians; 1*/η_L_* is the mean time to recapture.

| Re-isolation rate | Actual infections averted | Reduction (%) |
| --- | --- | --- |
| $\eta_L = 0.05 \text{ day}^{-1}$ ($\sim 20 \text{ d}$) | 49 | 4.4 |
| $\eta_L = 0.10 \text{ day}^{-1}$ ($\sim 10 \text{ d}$) | 99 | 8.9 |
| $\eta_L = 0.20 \text{ day}^{-1}$ ($\sim 5 \text{ d}$) | 158 | 14.1 |

**Table 4:** Averted actual infections when re-isolation (*η_L_* = 0.20 day*^−^*^1^) is switched on after intervention week *X*, relative to no re-isolation. Values are posterior medians.

| Intervention timing | Actual infections averted | Reduction (%) |
| --- | --- | --- |
| $\eta_L$ is 0.20 after ... | | |
| week 1 | 149 | 13.3 |
| week 2 | 106 | 9.4 |
| week 3 | 44 | 3.9 |
| week 4 | 13 | 1.1 |

Figure 7 (b) and Table 5 examine the impact of shortening the time from case detection to isolation by scaling the fitted rate *η̂* with *κ_η_*. In this context, 1*/η* denotes the average number of days until isolation. Among all levers considered, this one is clearly the most potent. Increasing *κ_η_* to 1.5 prevents 497 actual infections (44.4%), raising it to 2 prevents 684 (61.1%), and setting it to 3 prevents 828 (73.9%). In Figure 7 (b), all three observables decline sharply, with the trajectories diverging early, unlike the relatively modest and late-emerging effects of re-isolation shown in Figure 7 (a). This contrast arises from the underlying mechanism that isolation speed influences pre-isolation transmission for *every* detected case and shortens the period during which each contributes to the force of infection, whereas re-isolation and leakage control act only on the smaller subset of cases that escape. Collectively, these findings suggest that reducing the delay to isolation is the most influential control lever. Early leakage suppression still yields considerable, but time-limited, benefit, and re-isolation provides an additional, though secondary, advantage when complete prevention of escape cannot be achieved.

**Table 5:**
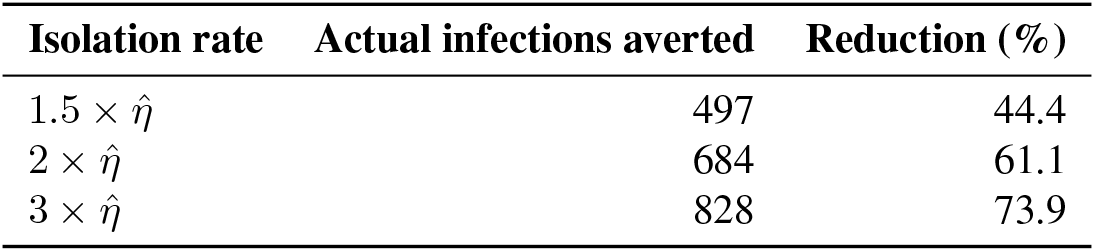
Averted actual infections under faster isolation of detected cases, relative to the fitted isolation rate *η̂*. Values are posterior medians; the scale factor multiplies *η̂*, shortening the mean pre-isolation window 1*/η*.

| Isolation rate | Actual infections averted | Reduction (%) |
| --- | --- | --- |
| $1.5 \times \hat{\eta}$ | 497 | 44.4 |
| $2 \times \hat{\eta}$ | 684 | 61.1 |
| $3 \times \hat{\eta}$ | 828 | 73.9 |

Figure 8 and Table 6 present the cumulative actual infections averted as each lever is increased from the fitted-model baseline to its maximum intensity. The maximum intensity is defined as *q_t_ ×* 3 for detection, *η ×* 3 for isolation speed, *η_L_* = 0.20 day*^−^*^1^ for re-isolation, *ρ* reduced to 0 for leakage, and *ϕ_D_* increased to 1 for safe burial. The averted infections are computed relative to the fitted model. The horizontal axis is scaled to each lever’s own range, so the comparison reflects each lever’s achievable maximum and the shape of its response curve (including diminishing returns). For curves that are truly concave, the “knee” indicates the intensity level beyond which additional effort produces strongly diminishing returns.

**Figure 8:**
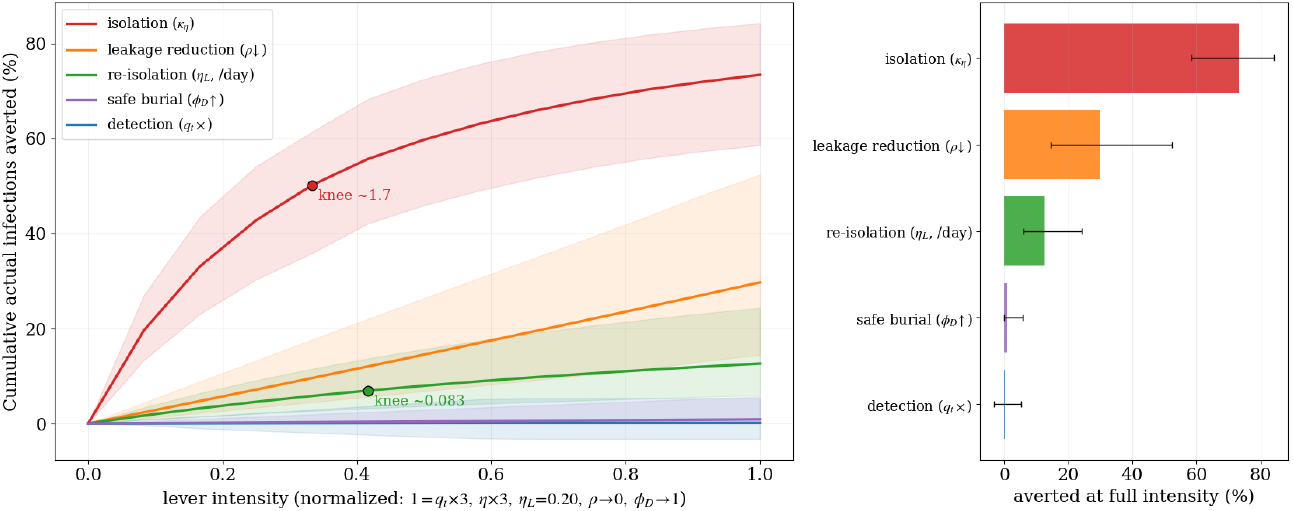
Knee analysis of the five intervention levers, its 95% HDI.

**Table 6:** Comparison of the five control levers by cumulative actual infections averted at full intensity, relative to the fitted-model baseline. Values are posterior medians with 95% HDIs in brackets. Full intensity corresponds to *q_t_ ×* 3, *η ×* 3, *η_L_* = 0.20 day*^−^*^1^, *ρ →* 0, and *ϕ_D_ →* 1; the axis therefore compares reachable ceilings and curvature within each lever’s plausible range, not equal operational effort. The knee (native units) marks the onset of diminishing returns (Kneedle), reported only where the curve is genuinely concave.

| Lever | Averted (%) | Knee (native) |
| --- | --- | --- |
| Isolation speed ($\kappa_\eta$) | 73.4 [59, 84] | $1.7 \times$ baseline value |
| Leakage reduction ($\rho \downarrow$) | 29.7 [14, 52] | near-linear |
| Re-isolation ($\eta_L$) | 12.6 [6, 24] | $0.083 \text{ day}^{-1}$ |
| Safe burial ($\phi_D \uparrow$) | 0.9 [0, 6] | near-linear |
| Detection ($q_t \times$) | 0.1 [-3, 5] | near-linear |

Isolation speed is by far the most influential lever. It achieves the highest impact, preventing 73.4% (95% HDI 59–84) of actual infections at *κ_η_* = 3. It is also the only lever that shows a clear knee in its response curve, occurring around *κ_η_* = 1.7. Past this point, further reductions in the pre-isolation period yield diminishing returns, implying that roughly halving the time to isolation captures most of the possible benefit. Leakage reduction ranks second in effectiveness, averting 29.7% (14–52) of infections as *ρ* approaches zero. Its response is nearly linear across the examined range, suggesting a consistent benefit without an obvious saturation threshold. Re-isolation has an intermediate effect, preventing 12.6% (6–24) of infections at *η_L_* = 0.20 day*^−^*^1^. Its curve exhibits a knee near 0.083 day*^−^*^1^, corresponding to a recapture time of about twelve days, beyond which faster recapture offers limited additional improvement.

The other two levers are weak across their reasonable ranges. Even at full coverage (*ϕ_D_* = 1), safe burial prevents only 0.9% (0–6) of infections. This aligns with the funeral transmission route accounting for only a small portion of the overall force of infection under the fitted model. Detection is essentially inactive, preventing just 0.1% (*−*3–5) of infections. The 95% HDI includes zero, indicating that intensifying contact follow-up does not materially reduce the actual number of infections. This may reflect the fact that better detection simply channels more cases into a still-leaky isolation system, in which detected and undetected individuals transmit at similar rates until they are isolated. Neither lever shows a meaningful knee, as both response curves remain essentially flat across their entire ranges.

Reducing the delay to isolation emerges as the single most potent and efficient lever, yielding most of its impact even at moderate levels of improvement. Eliminating leakage adds a substantial, steadily accumulating secondary benefit. Re-isolation provides a modest, intermediate advantage when complete prevention of escape is not feasible, while enhanced safe burial or improved detection, considered in isolation, are not sufficient to materially control the outbreak under the fitted parameters. We stress that these comparisons keep each lever’s plausible range fixed. Isolation speed is represented here as an idealized scaling factor. Whether a two- to three-fold acceleration in isolation is operationally achievable is a separate issue that lies outside the scope of this study.

## Discussion

Previous mathematical models of Ebola outbreaks have consistently highlighted the importance of separating transmission by mode.^13,12,8^ When community-based transmission is dominant, key interventions include early case detection, prompt isolation, measures to reduce risk within households, and systematic contact tracing. In contrast, for transmission associated with funerals, the main priorities are to ensure safe, dignified burials and to foster community participation in adapting safe traditional practices. If nosocomial transmission is involved, for instance, due to lapses in the use of personal protective equipment or overcrowded healthcare facilities, the emphasis shifts to infection prevention and the protection of healthcare workers.^13,12^ In the model described here, hospital-acquired transmission is intentionally assumed to be zero; patients in the hospital are considered completely and effectively isolated. We acknowledge that shortages of PPE and overcrowding in peripheral treatment units may still lead to some hospital-acquired exposures that are not accounted for by this assumption. This simplifying assumption is driven by our focus on examining how empirically observed isolation deficiency contributes to Ebola transmission and its control.

To our knowledge, very few modeling studies have explicitly incorporated isolation leakage as a distinct transmission route and estimated it jointly with under-detection during an ebolavirus outbreak.^7^ Our proposed SEUIHLF3R model extends the standard SEIHFR structure by introducing an escapee compartment *L* and an undetected community compartment *U* . By tying the leakage rate directly to observed provincial escapee counts and isolation census data, the SEUIHLF3R model yields a data-driven estimate of a control gap that is well recognized in operational reports but has not previously been quantified in a disease transmission framework. This helps fill an important gap in the literature. A recent systematic review reported that about 92% of Ebola virus disease models focus on the Zaire species^24^ and, to our knowledge, with little to no interest on leakage pathway, despite repeated accounts of escapees during previous Ebola outbreaks.^3,25,31^

Our main result is that isolation leakage is quantitatively significant, but its impact on outbreak size is highly time-dependent. Relative to the estimated baseline leakage rate, fully removing leakage could lower cumulative actual infections by about one-third, whereas doubling or tripling it leads to large increases. However, the advantage of reducing leakage erodes quickly with delay. Implementing strict control after one week yields roughly a 27% reduction in infections between 14th May and 17th June, but this falls to below 2% if implemented only by the fourth week. This timing pattern arises because leakage-driven transmission accumulates over time. By the later stages of the observation period, most transmission chains initiated by escapees are already in progress, so tightening leakage is mostly effective as an early intervention. Re-isolating patients who have already escaped provides an additional, albeit smaller, benefit, as it can only bring back individuals who have already left rather than prevent their escape in the first place. Importantly, keeping re-isolation in place from the beginning yields only slightly greater gains than simply eliminating leakage starting in the second week. Overall, the vast majority of the potential benefit from either approach is realized in the earliest stage of the response.

The decomposition of the reproduction number explains why detection and safe burial are relatively weak intervention levers under the fitted parameters. Since the model assumes that detected and undetected infections are equally infectious until isolation, raising detection primarily shifts the contribution to the overall reproduction number *R* from the undetected component *R_U_* to the pre-isolation component *R_I_*, rather than eliminating it. As a result, *R* would fall only slightly and would remain above one even with perfect detection. Safe burial affects only the funeral transmission route, which accounts for a relatively small fraction of the total infection pressure in this setting. This is further consistent with the relatively low case fatality and the sanitized treatment of deaths in hospitals and among detected community cases in the model. The control-boundary analysis translates these insights into an operational condition that dictates that successful control requires both adequate detection and sufficiently fast isolation. Put differently, there is a *detection floor* below which no isolation speed can bring *R* below one. This floor increases sharply with leakage, until, at around twice the fitted leakage rate, no practical combination of detection and isolation speed can achieve control. The main policy implication is that reducing the delay to isolation is the most potent single intervention lever, but it cannot compensate for poor leakage control once escape events become common.

These findings largely align with earlier Ebola modeling studies that highlight rapid case isolation and a shorter community infectious period as key drivers of epidemic magnitude.^13,12,2^ Our results extend these conclusions to a framework in which leakage is explicitly represented as a separate, empirically grounded process. The posterior *R*_0_ we obtain is somewhat higher than the CDC branching-process estimate for this outbreak, 3.67 vs. 2.51,^21^ which is unsurprising given the structural differences between the models. In our compartmental framework, early epidemic growth is ascribed to a combination of undetected community transmission, funeral-related transmission, and leakage, and *R*_0_ is interpreted as the pre-intervention transmissibility rather than an effective reproduction number that already incorporates control measures. The persistence of *R >* 1 across the detection and ascertainment parameter ranges further accords with the documented continued expansion of the Ituri epidemic over the analysis period.

Several limitations qualify the conclusions of this paper. First, the leakage pathway relies on reported escapee counts and the isolation census, both of which are likely under-recorded. As a result, the true magnitude of leakage may be underestimated. While the escapee likelihood partially accounts for this reliance on facility reporting, it does not eliminate it. In particular, escapes that occur at night and incidents that occur during security disruptions are unlikely to be recorded consistently.

Second, structural non-identifiability involving the undetected burden, the community fatality probability, the postmortem detection probability, and funeral infectiousness forced us to fix or strongly regularize several natural-history and funeral-process parameters, such as *T_U_*, *c_U_*, *ν*, and *m_F_* . The sensitivity analyses constrain, but do not eliminate, this dependence. In addition, the weak identifiability of *m_F_* and *ϕ_D_* in particular implies that the conclusions regarding the funeral pathway should be regarded as tentative. Besides, community deaths occurring in rural Ituri are not comprehensively recorded, implying that the unsafe-funeral compartment *F_U_*, and thus funeral-associated transmission, may be underestimated.

Third, we assume that undetected infections, pre-isolation infections, and infections arising from escapees have the same transmissibility, i.e., *β_U_* = *β_I_* = *β_L_*. If individuals who escape isolation, or symptomatic individuals prior to isolation, transmit at systematically higher or lower rates, the relative contribution of these routes would change. In particular, individuals who escape typically do so at later stages of illness. When symptoms are pronounced and infectiousness is highest, it is reasonable to assume that *β_L_ > β_U_* . This would increase, rather than decrease, the burden attributable to leakage, meaning that our estimates are conservative.

Fourth, reported proportions of contact follow-up may exaggerate the number of contacts truly reached in contexts of insecurity, displacement, and mistrust. Consequently, the high estimated detection probabilities (*p_d_ ≈* 0.91–0.99) should be interpreted as upper limits on the actual level of case ascertainment.

Fifth, the model does not incorporate spatial structure, age-stratified contact patterns, or explicit hospital-acquired transmission. Instead, it represents the Ituri epicenter as a single, homogeneously mixing population. In a context characterized by armed conflict and mass displacement, substantial heterogeneity in access to care and reporting is likely.

Sixth, the intervention analysis treats each control parameter as an idealized lever that can be adjusted independently. For example, the isolation-speed scenarios posit that a two- to three-fold increase in case isolation speed is achievable. Such efforts to speed up the response may be limited by insecurity, insufficient ambulance capacity, and community resistance. The intervention levers are evaluated over fixed, plausible parameter ranges rather than in terms of cost or operational feasibility. Consequently, the ranking captures epidemiological potential rather than public health practicality. Lastly, vaccination and therapeutics are not included in the model, consistent with the current lack of a licensed BDBV vaccine or approved treatment.^42,43,44^

From a public health perspective, our analysis supports a multilayered strategy that focuses on reducing delays in case isolation and on retaining patients in isolation centers. In conflict-affected, low-trust contexts such as Ituri, DRC, retention is not simply an operational challenge but is fundamentally driven by community engagement. Since people’s sense that isolation centers are unsafe leads them to flee, effective early engagement with communities and the establishment of trust are likely necessary conditions for achieving the leakage-control benefits identified here.

Our finding that leakage control is most beneficial in the early stages of an outbreak suggests that these investments should be concentrated up front. The detection-floor result also has a cautionary message for contexts where case detection cannot be raised above this threshold. In such settings, faster isolation alone is insufficient to halt transmission, making leakage reduction not discretionary but essential. This means that improving detection and isolation speed should happen together, not one after the other, particularly in contexts where insecurity constrains how fast detection can realistically be.

Future work should relax the assumptions of equal transmissibility and homogeneous mixing, incorporate spatial and care-access heterogeneity, and jointly model suspected and confirmed case trajectories to refine ascertainment estimates. Furthermore, as vaccines and therapeutics for BDBV become available, the framework should be expanded to assess pharmaceutical interventions alongside the non-pharmaceutical measures analyzed here.

## Data Availability

All data produced are available online at https://github.com/tamfatkh/ebola

## Author Contributions

T.O.: Conceptualization, Data Curation, Formal Analysis, Methodology, Software, Validation, Visualization, Writing – Original Draft Preparation, Writing – Review & Editing

D.F.: Conceptualization, Writing – Review & Editing

M.L.N-M.: Conceptualization, Methodology, Writing – Original Draft Preparation, Writing – Review & Editing

All authors reviewed and approved the final manuscript.

## Ethics Statement

This study uses aggregated publicly available surveillance data and an officially released situation report from the Institut National de Santé Publique of the DRC. No individual patient data were accessed. No ethics committee approval was required for this secondary analysis of aggregate surveillance data.

## Declaration of generative AI and AI-assisted technologies in the manuscript preparation process

During the preparation of this work, the author(s) used Claude and Grammarly to edit and correct the English language. After using these tools, the author(s) reviewed and edited the content as needed and took full responsibility for the content of the published article.

## Acknowledgements

The authors thank the COUSP/SGI response team and the Institut National de Santé Publique of the DRC for making facility-level surveillance data available. The authors acknowledge the health workers and responders in Ituri Province operating under severe conditions during the 17th DRC Ebola epidemic.

## S1. Supplementary Material

### S1.1. Outbreak Data

**Table S1:** Provincial situation-report series for the 17th DRC Ebola epidemic, 14 May-17 June 2026 (Ituri Province epicenter). Escapees are patients who left isolation without being discharged. The isolation census and contact-follow-up rate are reported from 30 May / 1 June onward. CFR is deaths as a percentage of confirmed cases. Dashes denote quantities not reported in that situation report.

| N° | Date | Conf. cases | Deaths | Discharged | In isol. | Escapees | F/U % | CFR % |
| --- | --- | --- | --- | --- | --- | --- | --- | --- |
| 001 | 14 May | 8 | 4 | - | - | - | - | 50.0 |
| 002 | 17 May | 13 | - | 0 | - | - | - | - |
| 004 | 19 May | 33 | 4 | - | - | - | - | 12.1 |
| 005 | 19 May | 51 | 4 | 0 | - | - | - | 7.8 |
| 006 | 20 May | 64 | 6 | 0 | - | 4 | - | 9.4 |
| 007 | 21 May | 83 | 9 | 0 | - | 10 | - | 10.8 |
| 008 | 22 May | 91 | 10 | 0 | - | - | - | 11.0 |
| 009 | 23 May | 101 | 10 | 0 | - | 23 | - | 9.9 |
| 010 | 24 May | 105 | 10 | 0 | - | 6 | - | 9.5 |
| 011 | 25 May | 106 | 12 | 0 | - | - | - | 11.3 |
| 012 | 26 May | 121 | 17 | 0 | - | - | - | 14.0 |
| 013 | 27 May | 125 | 17 | 1 | - | 4 | - | 13.6 |
| 014 | 28 May | 210 | 17 | 1 | - | 4 | - | 8.1 |
| 015 | 29 May | 263 | 42 | 2 | - | 15 | - | 16.0 |
| 016 | 30 May | 282 | 42 | 2 | - | 5 | 45.0 | 14.9 |
| 017 | 31 May | 321 | 48 | 6 | - | 7 | - | 15.0 |
| 018 | 01 Jun | 344 | 60 | 6 | 173 | 4 | 43.0 | 17.4 |
| 019 | 02 Jun | 363 | 62 | 6 | 206 | 11 | 45.0 | 17.1 |
| 020 | 03 Jun | 381 | 64 | 7 | 233 | 20 | 55.0 | 16.8 |
| 022 | 05 Jun | 488 | 86 | - | 267 | 10 | 67.0 | 17.6 |
| 023 | 06 Jun | 515 | 91 | - | 283 | 6 | 50.0 | 17.7 |
| 024 | 07 Jun | 550 | 101 | - | 309 | 10 | 64.0 | 18.4 |
| 025 | 08 Jun | 598 | 115 | - | 297 | 12 | 56.0 | 19.2 |
| 026 | 09 Jun | 635 | 127 | 30 | 260 | 9 | 61.0 | 20.0 |
| 027 | 10 Jun | 676 | 136 | 32 | 262 | 3 | 71.0 | 20.1 |
| 028 | 11 Jun | 689 | 139 | 32 | 315 | 4 | 28.0 | 20.2 |
| 030 | 13 Jun | 782 | 181 | 40 | 359 | 0 | 56.0 | 23.1 |
| 031 | 14 Jun | 808 | 192 | 48 | 363 | 1 | 63.0 | 23.8 |
| 032 | 15 Jun | 837 | 196 | 49 | 376 | 6 | 64.0 | 23.4 |
| 033 | 16 Jun | 875 | 202 | 67 | 379 | 1 | 71.0 | 23.1 |
| 034 | 17 Jun | 896 | 232 | 78 | 383 | 0 | 71.0 | 25.9 |
| Total escapees reported |  |  |  |  |  | 175 |  |  |

#### S1.2. Modeling Assumptions

Table S2 presents the primary modeling assumptions, their intended purposes, and the implications for interpretation.

**Table S2:**
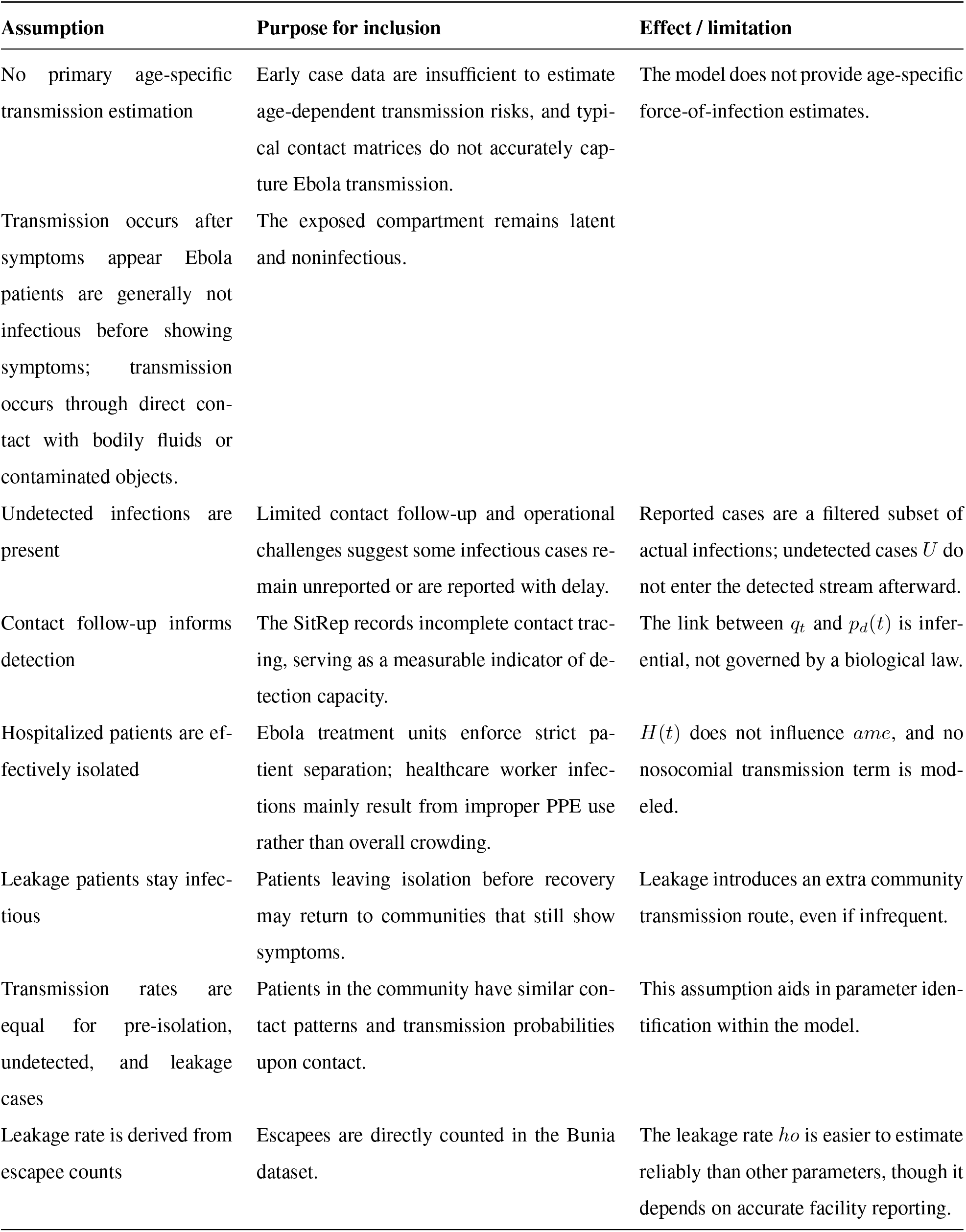
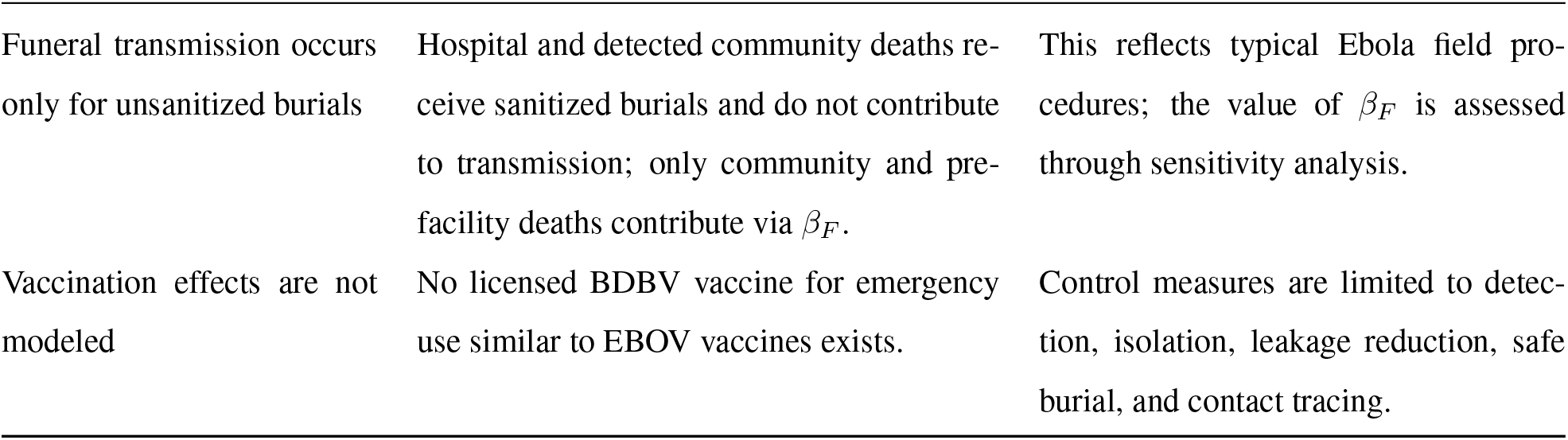
Summary of key modeling assumptions and their effects.

| Assumption | Purpose for inclusion | Effect / limitation |
| --- | --- | --- |
| No primary age-specific transmission estimation | Early case data are insufficient to estimate age-dependent transmission risks, and typical contact matrices do not accurately capture Ebola transmission. | The model does not provide age-specific force-of-infection estimates. |
| Transmission occurs after symptoms appear Ebola patients are generally not infectious before showing symptoms; transmission occurs through direct contact with bodily fluids or contaminated objects. | The exposed compartment remains latent and noninfectious. |  |
| Undetected infections are present | Limited contact follow-up and operational challenges suggest some infectious cases remain unreported or are reported with delay. | Reported cases are a filtered subset of actual infections; undetected cases $U$ do not enter the detected stream afterward. |
| Contact follow-up informs detection | The SitRep records incomplete contact tracing, serving as a measurable indicator of detection capacity. | The link between $q_t$ and $p_d(t)$ is inferential, not governed by a biological law. |
| Hospitalized patients are effectively isolated | Ebola treatment units enforce strict patient separation; healthcare worker infections mainly result from improper PPE use rather than overall crowding. | $H(t)$ does not influence $ame$, and no nosocomial transmission term is modeled. |
| Leakage patients stay infectious | Patients leaving isolation before recovery may return to communities that still show symptoms. | Leakage introduces an extra community transmission route, even if infrequent. |
| Transmission rates are equal for pre-isolation, undetected, and leakage cases | Patients in the community have similar contact patterns and transmission probabilities upon contact. | This assumption aids in parameter identification within the model. |
| Leakage rate is derived from escapee counts | Escapees are directly counted in the Bunia dataset. | The leakage rate $ho$ is easier to estimate reliably than other parameters, though it depends on accurate facility reporting. |
| Funeral transmission occurs only for unsanitized burials | Hospital and detected community deaths receive sanitized burials and do not contribute to transmission; only community and pre-facility deaths contribute via $\beta_F$. | This reflects typical Ebola field procedures; the value of $\beta_F$ is assessed through sensitivity analysis. |
| Vaccination effects are not modeled | No licensed BDBV vaccine for emergency use similar to EBOV vaccines exists. | Control measures are limited to detection, isolation, leakage reduction, safe burial, and contact tracing. |

### S1.3. Model Parameters and Prior Distributions

BDBV-specific evidence is prioritized over all-species pooled estimates when determining the incubation period. A BDBV subgroup random-effects mean incubation period is estimated at 7.65 days (95% CI 6.43–8.88).^24^ Based on a sample of *n* = 116,^37^ the central estimated range across BDBV studies is 5.7–11.3 days.^37^ The baseline value of *σ* = 1*/*7.65 *≈* 0.131 day*^−^*^1^ is anchored to this BDBV-specific pooled estimate, with sensitivity analyses conducted over a range from 1/11.3 to 1/5.7 day*^−^*^1^. For comparison, MacNeil et al.^17^ reported mean incubation periods of 5.7 days among survivors and 7.4 days among fatal cases during the 2007 Uganda BDBV outbreak.

Statistically significant species differences in symptom-onset-to-recovery intervals have been reported (*p <* 0.01).^24^ The BDBV subgroup random-effects mean is 10.77 days (95% CI 9.74-11.80), whereas the all-species pooled estimate is 13.0 days (95% CI 10.4-15.7).^24,29,37^ The baseline *T_H_*= 10.8 days for in-isolation recovery is based on the BDBV-specific estimate, and the all-species estimate serves as the upper bound in sensitivity analyses.

The random-effects mean duration from symptom onset to death for the BDBV subgroup is 10.84 days (95% CI, 9.91-11.78), whereas the pooled estimate across all species is 9.3 days (95% CI, 8.5-10.1).^24,29,17,37^ The baseline time-to-death component uses the BDBV-specific estimate of 10.8 days.

The pooled random-effects mean infectious period across species is 5.0 days (95% CI 3.7–6.3; common-effects 5.41 days), based on three estimates from Zaire species studies. No BDBV-specific estimate of the infectious period is available in the literature.^24^ Accordingly, the community infectious duration *T_U_* is set to 5.0–5.4 days, using this all-species estimate as a Zaire-derived approximation for BDBV.

Table S3 presents the baseline parameter assignments and prior distributions. Rates are reported per day. The prior distributions are conservatively specified, given the limited identifiability of several biological parameters.

**Table S3:**
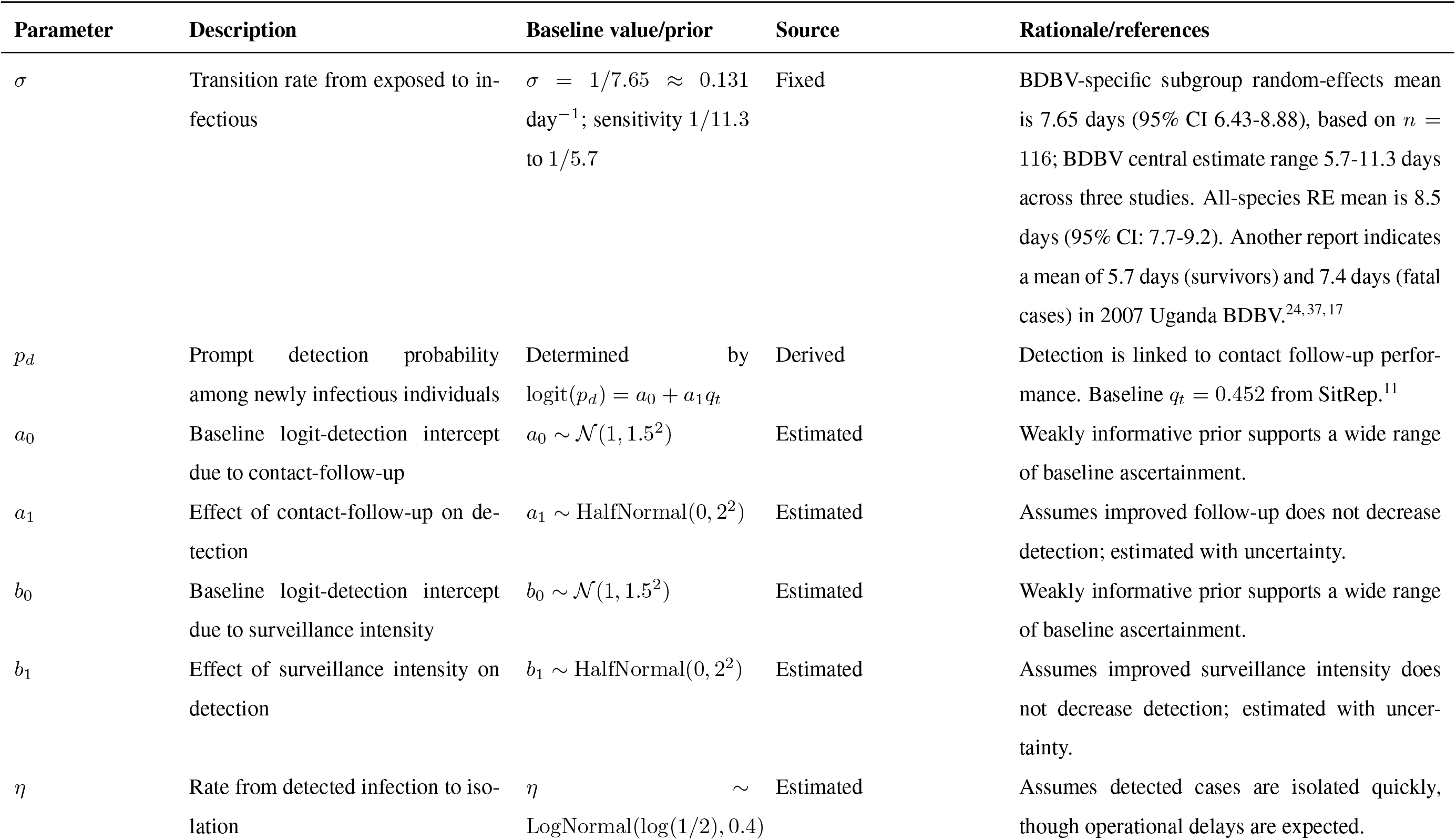
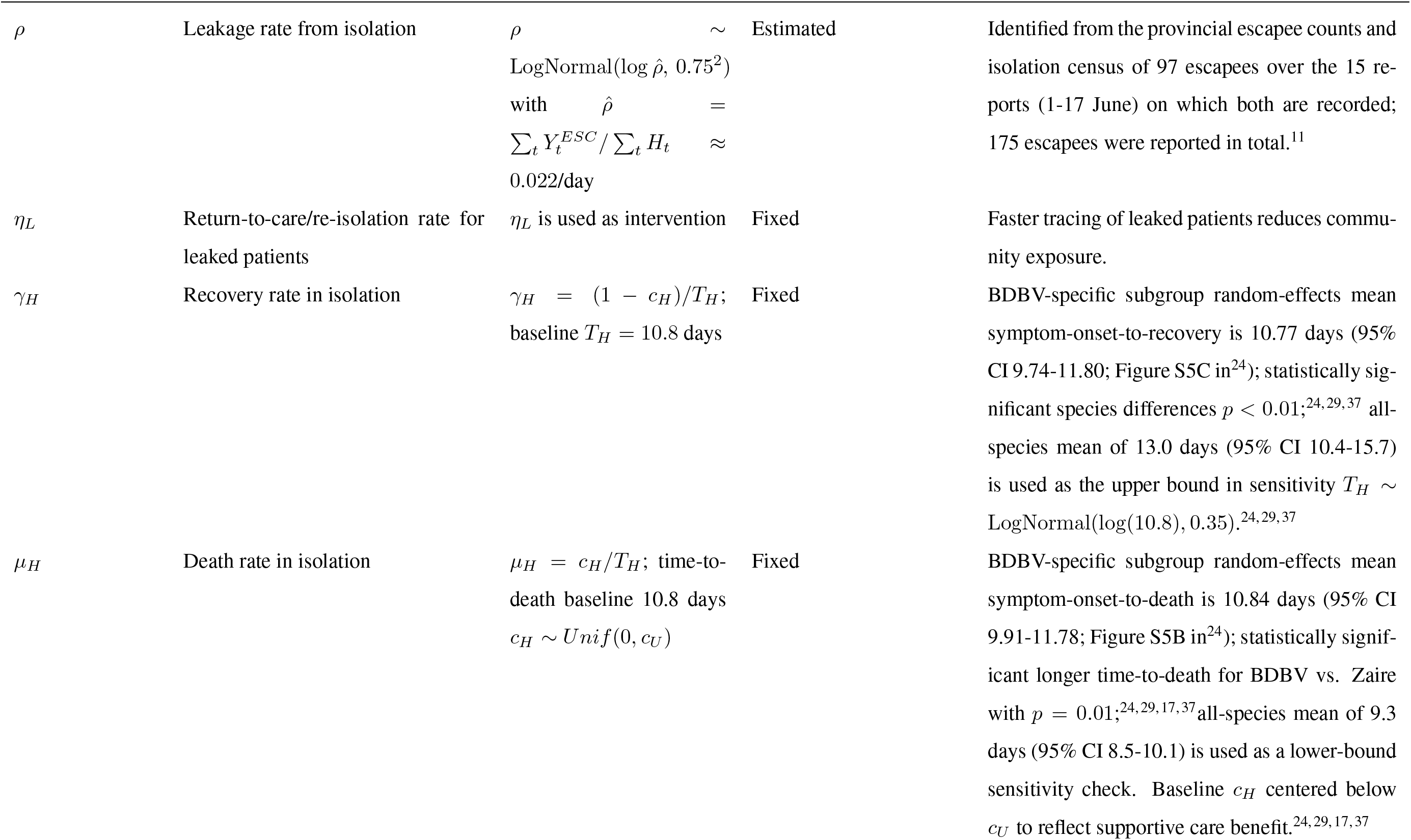
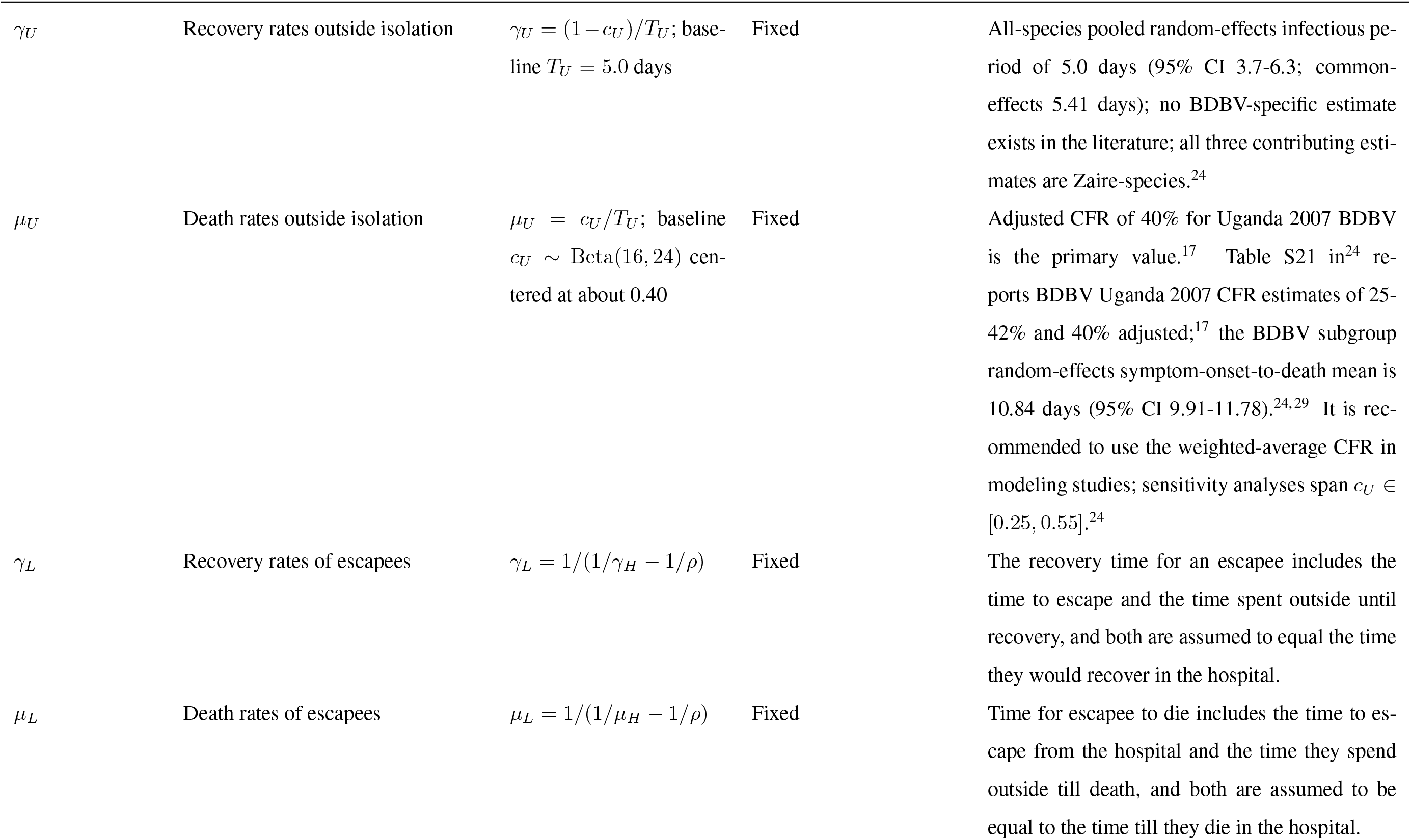
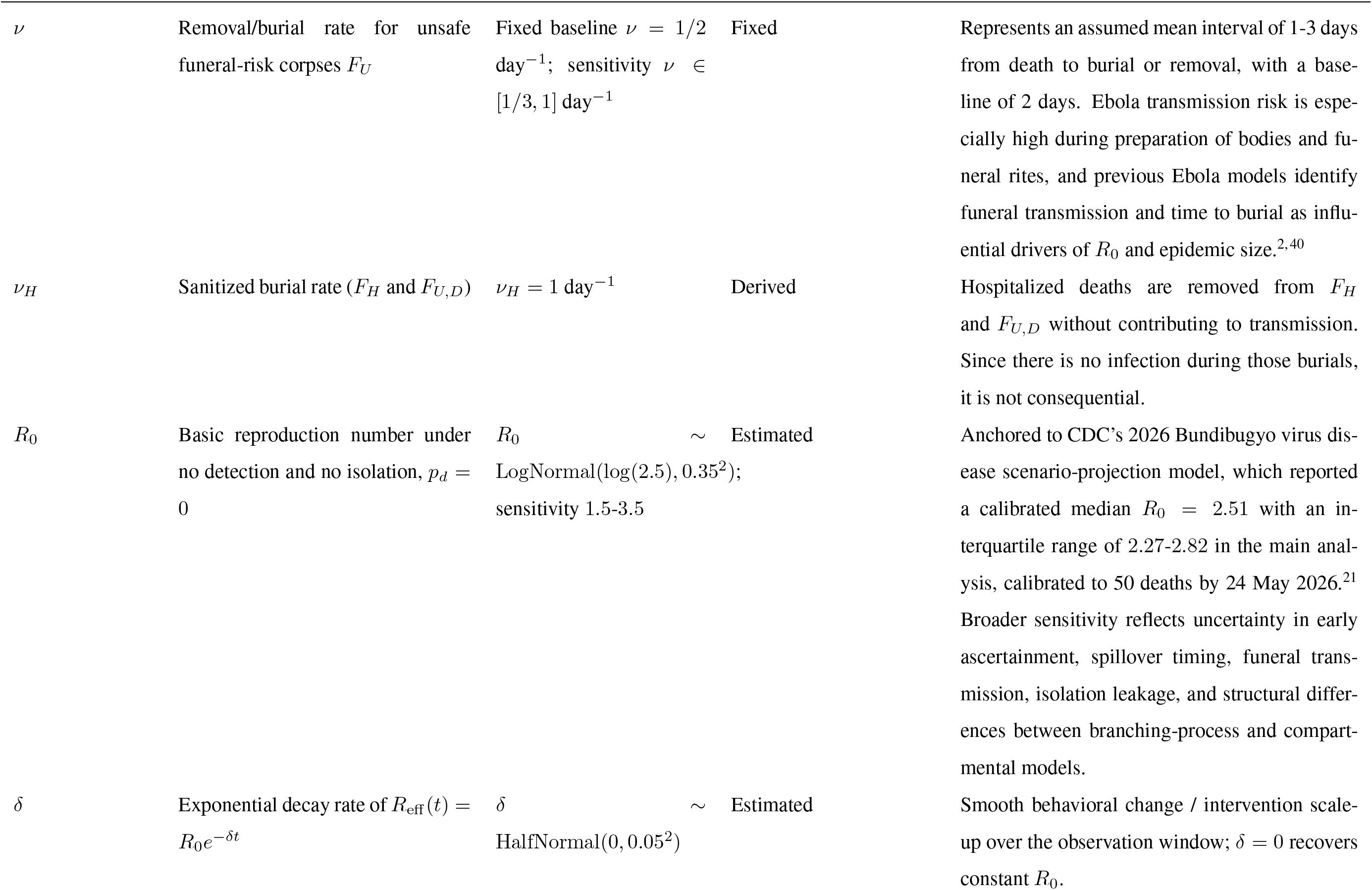
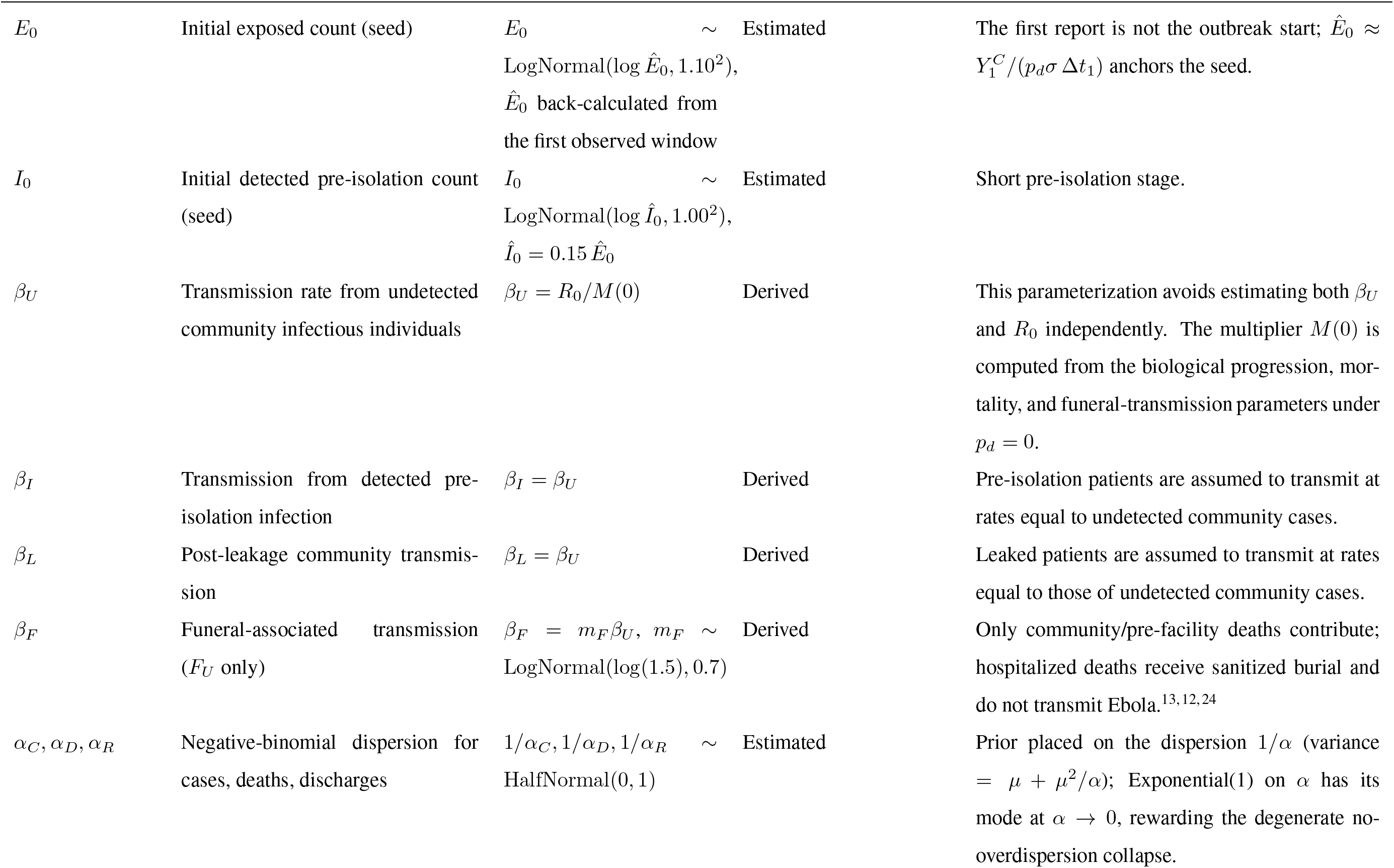
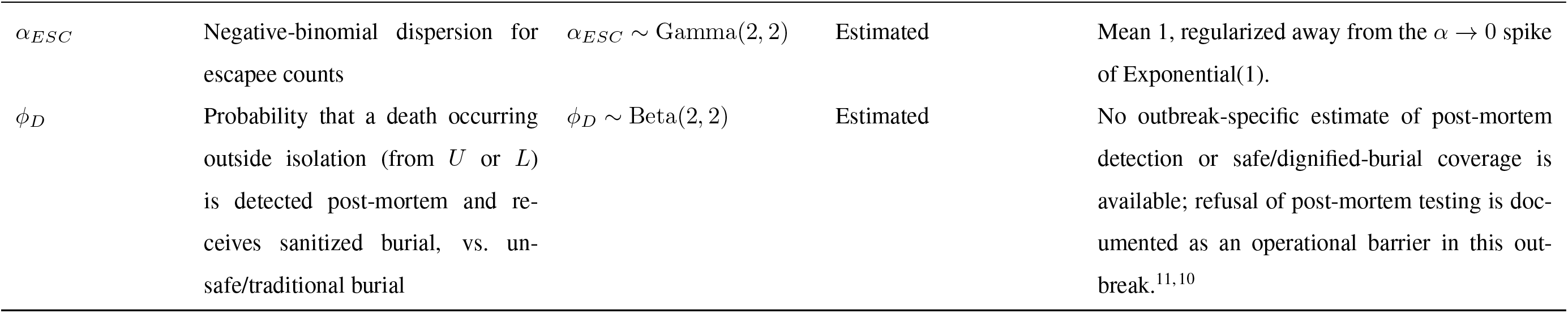
Baseline parameter values and prior distributions for the detection-isolation-leakage SEUIHLF3R model.

### S1.4. MCMC Outputs

**Figure S1:**
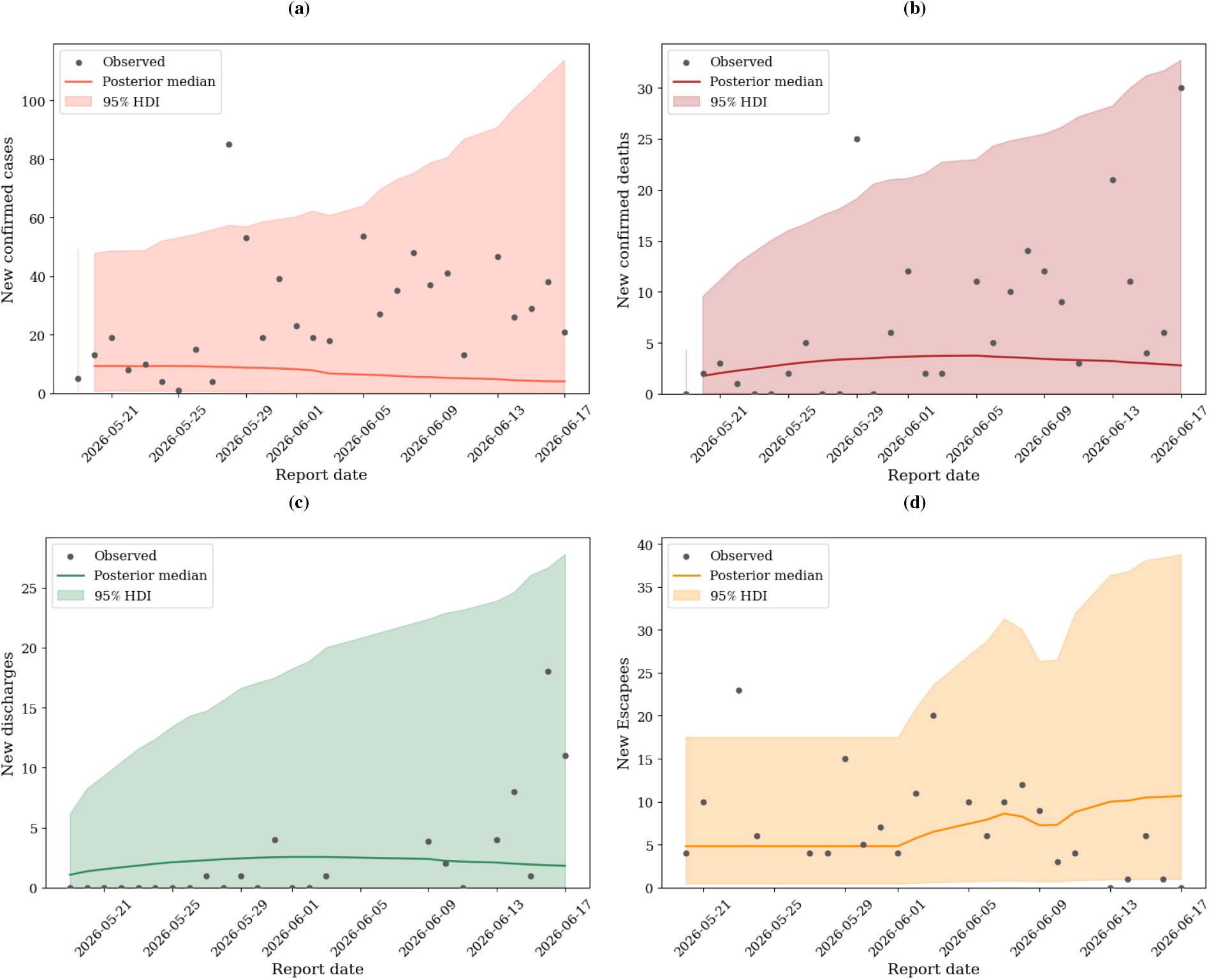
Median and 95% HDI of the prior predictive trajectories versus reported data of (a) the number of Ebola cases, (b) the number of deaths, (c) the number of recoveries, and (d) the number of escapees in Ituri province.

**Figure S2:**
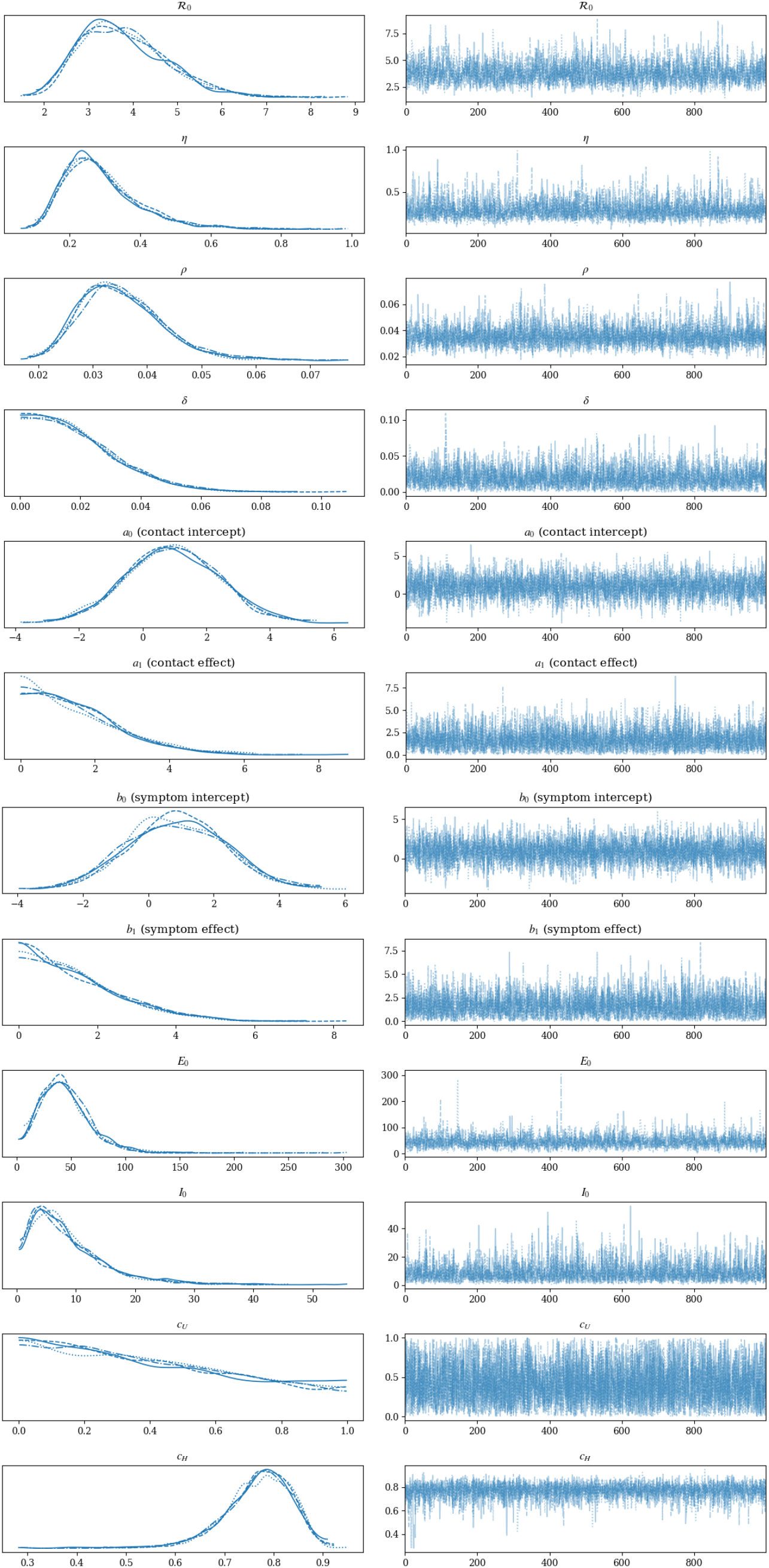
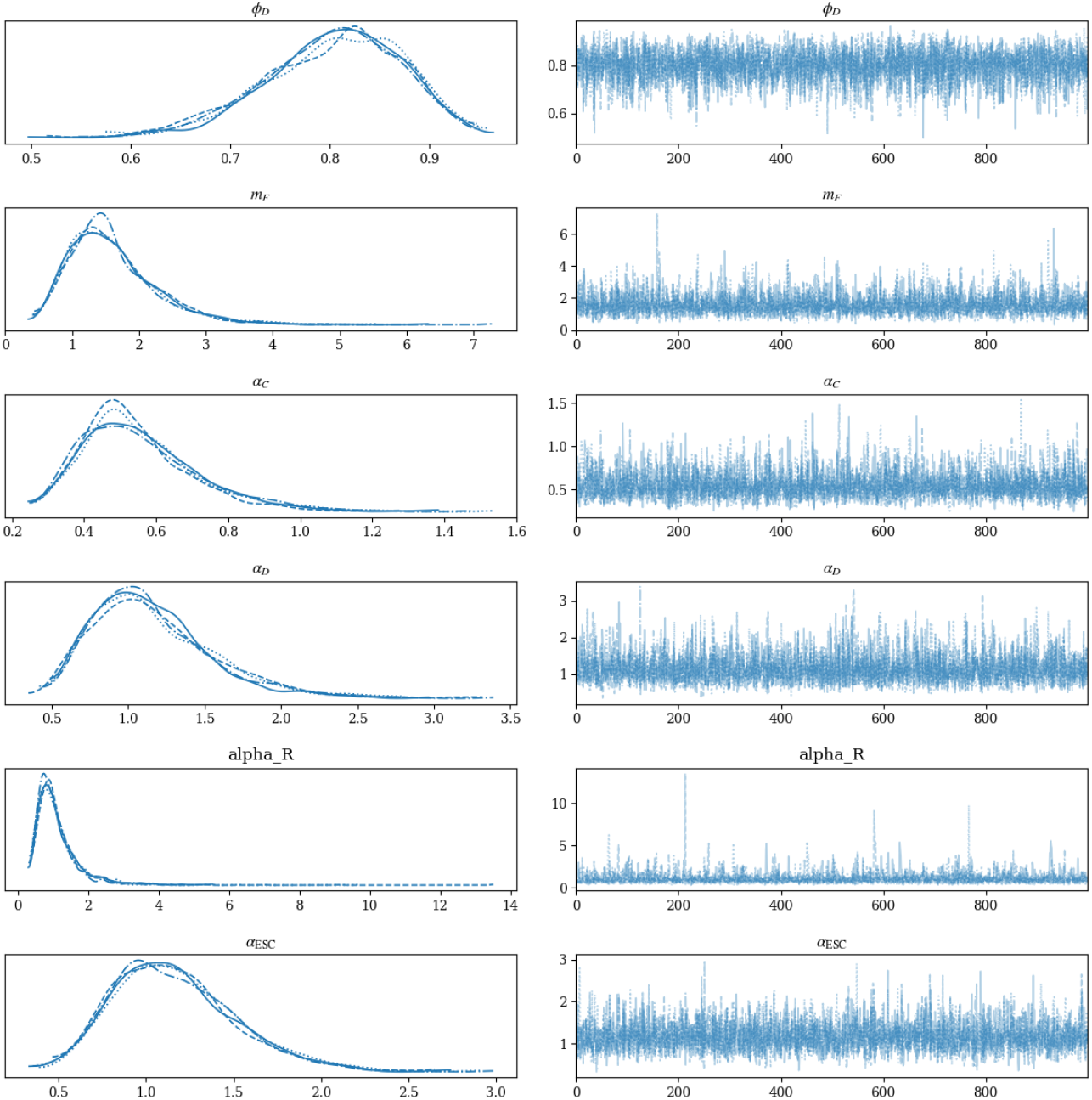
MCMC traces and posterior distributions of the estimated parameters indicate convergence of the four independent MCMC chains after the burn-in period of 1000 runs and 1000 retained runs. The MCMC’s started from four randomly selected initial parameter vectors.

**Figure S3:**
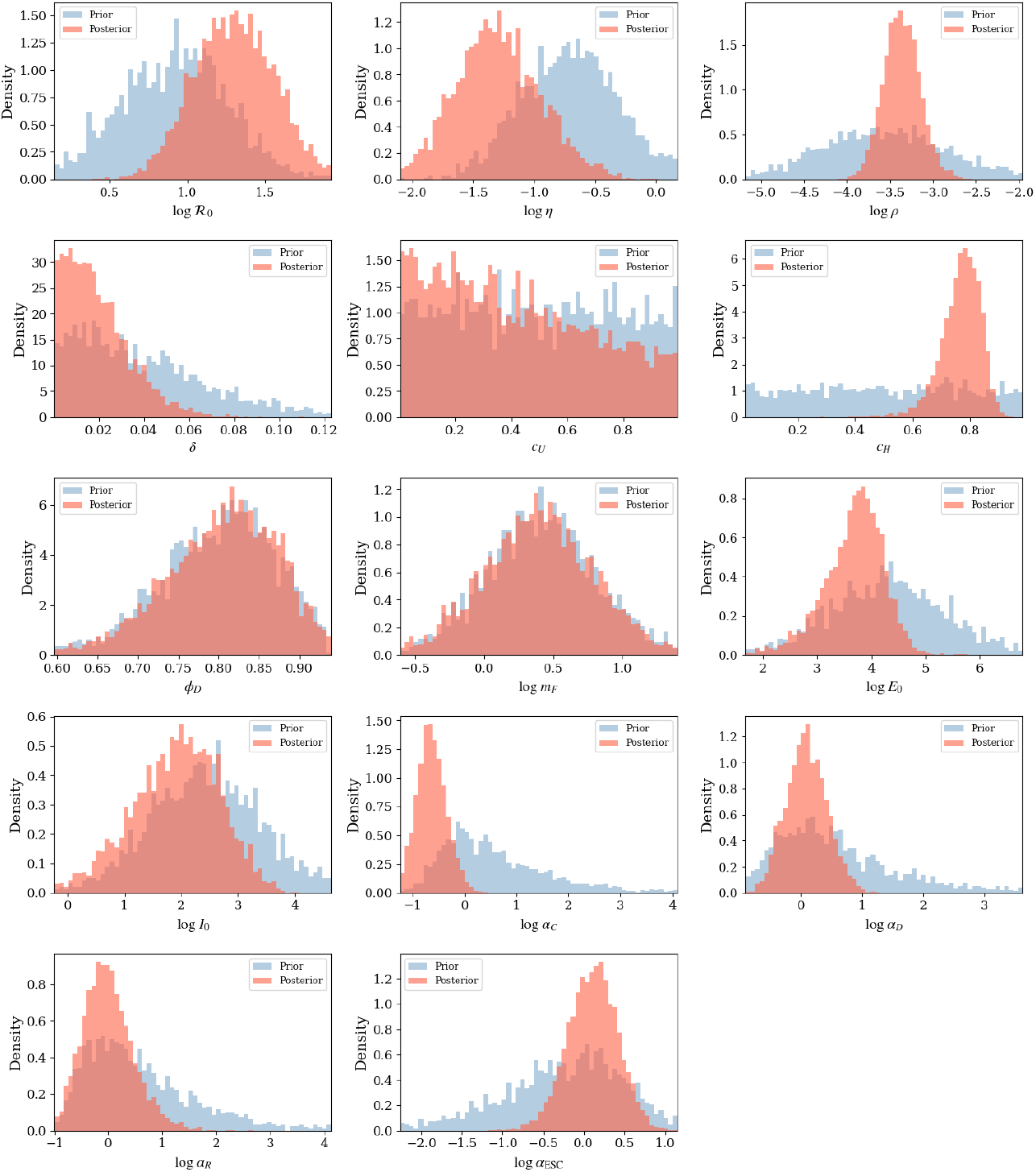
Posterior shifts and contractions from prior distributions of the parameters indicate that the model learned those parameters values from the data.

**Table S4:** Posterior summary statistics (95% HDI). *r̂* < 1.01 and ESS_bulk_, ESS_tail_ > 400 indicate adequate convergence. The results are based on four independent MCMC runs, each with a 1000-iteration burn-in and 1000 iterations.

| | Mean | Median | SD | HDI<sub>2.5%</sub> | HDI<sub>97.5%</sub> | $\hat{r}$ | $\text{ESS}_{\text{bulk}}$ | $\text{ESS}_{\text{tail}}$ |
| --- | --- | --- | --- | --- | --- | --- | --- | --- |
| $R_0$ | 3.804 | 3.672 | 1.001 | 1.982 | 5.675 | 1.001 | 1905 | 2300 |
| $\eta$ | 0.291 | 0.270 | 0.108 | 0.124 | 0.513 | 1.001 | 2494 | 2852 |
| $\rho$ | 0.035 | 0.034 | 0.008 | 0.022 | 0.051 | 1.001 | 3877 | 2436 |
| $\delta$ | 0.019 | 0.016 | 0.014 | 0.000 | 0.046 | 1.000 | 2338 | 1806 |
| $a_0$ (contact intercept) | 0.953 | 0.933 | 1.449 | -2.114 | 3.583 | 1.000 | 3928 | 3042 |
| $a_1$ (contact effect) | 1.584 | 1.344 | 1.210 | 0.000 | 3.971 | 1.002 | 3702 | 1861 |
| $b_0$ (symptom intercept) | 0.890 | 0.876 | 1.479 | -2.154 | 3.634 | 1.003 | 5023 | 3160 |
| $b_1$ (symptom effect) | 1.517 | 1.253 | 1.193 | 0.002 | 3.849 | 1.001 | 2658 | 1634 |
| $E_0$ | 45.011 | 42.014 | 23.566 | 6.440 | 88.284 | 1.001 | 1958 | 1899 |
| $I_0$ | 8.708 | 7.018 | 6.530 | 0.578 | 21.976 | 1.001 | 2821 | 2687 |
| $c_U$ | 0.416 | 0.379 | 0.278 | 0.001 | 0.916 | 1.000 | 4231 | 2376 |
| $c_H$ | 0.768 | 0.777 | 0.070 | 0.634 | 0.889 | 1.002 | 3806 | 2605 |
| $\phi_D$ | 0.803 | 0.810 | 0.069 | 0.675 | 0.937 | 1.001 | 4219 | 2637 |
| $m_F$ | 1.602 | 1.481 | 0.665 | 0.555 | 2.931 | 1.001 | 4851 | 2918 |
| $\alpha_C$ | 0.553 | 0.523 | 0.162 | 0.298 | 0.888 | 1.003 | 4018 | 2546 |
| $\alpha_D$ | 1.160 | 1.092 | 0.400 | 0.497 | 1.938 | 1.002 | 4386 | 2928 |
| $\alpha_R$ | 1.124 | 0.957 | 0.689 | 0.350 | 2.370 | 1.002 | 3173 | 2387 |

**Figure S4:**
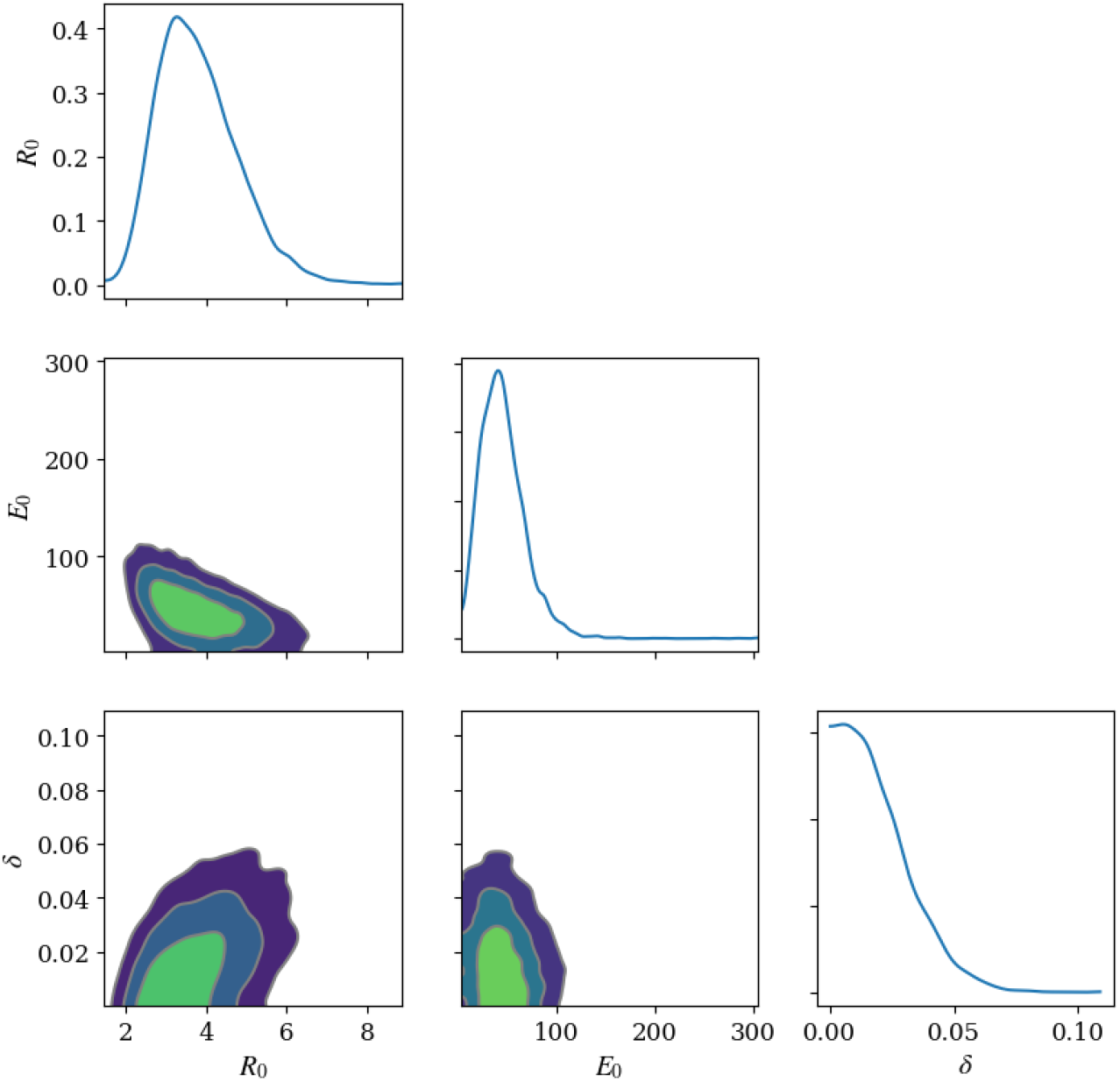
Identifability check up.

